# Think-HF: Development, feasibility, and usability of an electronic health record integrated tool for identifying missed diagnoses of heart failure in primary care

**DOI:** 10.64898/2026.01.23.26344713

**Authors:** K Barber, C Deaton, GP McCann, L Bernhardt, S Prinjha, R Alaei Kalajahi, MR Ali, I Squire, CJ Taylor, JGF Cleland, K Khunti, C Lawson

## Abstract

**Background:** Many patients with heart failure (HF) remain undiagnosed until acute hospital admission for decompensation. Think-HF is a clinical decision support tool (CDST) designed to identify possible undiagnosed HF in primary care and trigger timely, guideline recommended assessment.

**Methods:** Think-HF was developed through epidemiological evidence and codesign. Integrated within SystmOne, it analyses routine primary care data (codes, medications, test results, free text) to detect missed or emerging HF signals. When a record is opened, if a patient has ≥ two comorbidity indicators plus an HF-suggestive symptom, an alert is triggered and a structured template opens with one-click options for natriuretic peptide (NP) testing, echocardiography, specialist referral and coding. Additional algorithms identify unresolved investigations, coding inconsistencies and medication-based signals, generating a marker on the patient’s record. A mixed-methods feasibility study across six primary care practices assessed reach, usability, acceptability, and early implementation signals, using the RE-AIM framework.

**Results:** Six socioeconomically and ethnically diverse general practices participated (63 GPs; 78,640 patients [47.5% women, 59.4% White, 17.8% South Asian, 8.5% Black, 3% Chinese and 2.8% mixed ethnicities]). At baseline, 876 patients (1.1%) met the main alert criteria and 2,805 (3.6%) met additional algorithm criteria. Of 801 patients on the HF register, 665 (83%) lacked a refined HF left ventricular phenotype code. During four months of testing, Think-HF generated 299 clinically relevant main trigger alerts (75% during consultations). Early improvements included (i) 31 new HF diagnoses (+4.2%), with higher gains (+10%) in practices with lower baseline prevalence; (ii) a 14% increase in refined HF phenotype coding; (iii) fewer raised NT-proBNP results without follow-up (−7%); and (iv) fewer patients prescribed loop diuretics without NP testing (−14%, up to −45% in one practice with pharmacist-supported review). Clinicians reported improved awareness, more systematic assessment, and better follow-up of missed investigations or coding anomalies. Identified gaps aligned with patient-reported delays and misattributed symptoms.

**Conclusions:** Think-HF is feasible, acceptable and well aligned with routine primary care workflows. Early gains in diagnostic processes and coding accuracy highlight its potential to improve patient care. Team-based implementation will be essential for scale-up. Larger evaluation is required to assess clinical impact.

## Introduction

Heart failure (HF) is a leading cause of preventable hospital admissions in Europe.(1) Much of this burden reflects missed opportunities for earlier diagnosis, as many patients are only identified once critically unwell and needing emergency care.(2) This late recognition has serious consequences, with many patients presenting in acute decompensation and a minority dying before diagnosis is ever made.(3,4) Outcomes are considerably worse for those diagnosed in hospital rather than in primary care,(5–7) yet early identification in general practice remains challenging. Patients often present with non-specific symptoms,(8,9) particularly in HF with preserved ejection fraction, (10,11) or multiple long-term conditions (multimorbidity) creating uncertainty for both patients(8) and clinicians.(12) Because HF is defined by symptoms, signs, and objective evidence of cardiac dysfunction,(13) diagnosis depends on clinical suspicion, and timely investigation.

Although guidelines recommend natriuretic peptide testing followed by echocardiography and specialist review where appropriate,(14,15) many patients later diagnosed with HF do not follow this pathway. Diagnostic delays are common, and substantial variation in care persists.(16–18) Around two-thirds of patients have early indicators of HF in primary care, yet recommended tests are underused and approximately half undergo no diagnostic testing before diagnosis.(2,19) These shortfalls reflect clinical and system-level barriers,(20) including variable access to diagnostics, inconsistent implementation of guidance, and non-specific coding.(21,22) As a result, patients may not be added to HF registers and can miss guideline-recommended therapy and ongoing monitoring. Identifying those at highest risk of undiagnosed HF in primary care is therefore critical to optimise use of diagnostic resources, reduce avoidable hospitalisations, and improve outcomes.

Computerised decision support tools (CDSTs) offer a solution by providing real-time, patient-specific prompts that support clinical decision-making. CDSTs improve practitioner performance and patient outcomes across prevention,(23) diagnosis,(24) prescribing,(25,26) and guideline implementation.(27) However, simple ‘information-only’ alerts risk fatigue; effective systems require seamless integration, tailored recommendations, and actionable support. When these features are combined, CDSTs improved clinical practice in 94% of 32 randomised trials.(28) In PROMPT-HF, electronic tailored assessments increased guideline-recommended HF prescribing by 40% in outpatient clinics.(29) Building on this evidence, we developed Think-HF, a primary care-focused CDST co-designed with PRIMIS to support earlier recognition and diagnosis of HF. Think-HF aligns with the NHS 10-Year Plan by supporting proactive, digitally enabled HF care in the community.(30) This paper describes its development and pilot testing in six general practices

## Methods

### Phase 1: Tool Development

The development process followed four sequential stages; the first two have been published previously and are summarised here for completeness.

#### Stage 1-2: Evidence Base and Indicator Development

In brief, a large epidemiological analysis of 1,662 general practices demonstrated substantial missed opportunities for earlier HF recognition and underuse of diagnostic pathways, particularly among patients with early symptoms or proxy indicators such as loop diuretics.(19) These findings, alongside existing quantitative and qualitative research,(5,8,9,11,12,31–34) stakeholder consensus work,(35) and national patient survey insights,(36) informed a long-list of clinical and system-level indicators of possible undiagnosed HF. A modified Delphi process involving clinicians across primary and secondary care and individuals with lived experience prioritised indicators most suitable for electronic identification.(37) High-priority indicators included unresolved investigations (e.g., raised NT-proBNP without referral), medication-based indicators (particularly loop diuretics), and relevant comorbidity patterns. These outputs formed the basis for subsequent refinement and operationalisation.

#### Stage 3: Workflow Observation and Contextual Integration

To understand how Think-HF could operate within real primary care workflows, structured observations of routine clinical consultations were conducted across three general practices. GPs assessing patients presenting with symptoms were observed during routine clinical consultations. The clinicians and patients were asked for their consent for the consultation to be observed by a research nurse. A brief practice questionnaire and standardised observation template captured contextual factors, clinical behaviours, consultation flow, and potential integration points for CDST use (**S1 methods**).

#### Stage 4: Indicator Refinement and Operationalisation for Algorithm Development

The candidate indicators were refined iteratively to ensure relevance, clarity and feasibility for automated detection within GP electronic health records. Clinicians recruited from national heart failure clinical networks assessed each indicator using a Red-Amber-Green (RAG) approach, considering sensitivity, specificity and primary care applicability (**S2 Methods**). A second round focused on refining operational definitions and identifying any missing indicators.

Following indicator agreement and definition finalisation, the Think-HF algorithm and user interface was developed with PRIMIS, a specialist team that provides tools and training to support intelligent use of primary care data and to drive behavioural change in general practice.

### Phase 2: Feasibility Study Design

The feasibility phase assessed early implementation, usability, acceptability, fidelity and perceived sustainability of Think-HF in routine primary care. Findings were structured using the Reach, Effectiveness, Adoption, Implementation, Maintenance (RE-AIM) framework,(38) with a focus on implementation rather than clinical effectiveness. RE-AIM is a commonly used planning and evaluation framework across public health, behavioural science, and implementation science and aligns with key themes from the Medical Research Council Process Evaluation framework.(39)

Three sequential steps were undertaken:

1. Two-practice pilot - to assess early usability, acceptability, workflow fit and identify technical or operational refinements.
2. Refinement cycle - modification of the tool, definitions, supporting training or implementation materials.
3. Expanded testing in four additional practices - to explore variability in adoption, fidelity, usability and contextual barriers/enablers across a range of settings.

Each general practice used the tool for four months and process evaluation feedback was provided via baseline and follow-up surveys and post-intervention interviews with participating GPs. A sample of patients identified by Think-HF as having risk indicators for HF were also invited to interview to explore challenges in seeking assessment, navigating diagnostic pathways and engaging with care. These patients were not necessarily exposed to the tool; rather, their perspectives offered complementary insights into barriers, delays and opportunities for earlier recognition to inform future optimisation and wider implementation of Think-HF.

### Study Setting and Participating Practices

General practices in West Leicestershire, Leicester City and East Leicestershire and Rutland were invited to participate. Practices were purposively sampled to ensure inclusion across socioeconomically diverse areas.

*Eligibility Criteria Practice eligibility:*

- A list size of ≥ 6500 patients
- Use of SystmOne clinical IT system.
- Willingness to install and test Think-HF
- Agreement to participate in evaluation activities

*Individual clinician and patient eligibility for the process evaluation:*

- GPs involved in the assessment of patients
- Patients presenting to the GP practice for a consultation who are identified by Think-HF as having risk indicators for heart failure. Patients lacking mental capacity or in the dying phase of heart failure were excluded.

### Recruitment and Sampling Strategy

GP practices were identified and screened by the East Midlands National Institute for Health and Care Research (NIHR) Regional Research Delivery Network. Eligible practices were contacted via an invitation email containing study information. Practice managers and clinicians were offered a follow-up meeting (face-to-face or virtual) to discuss participation. Across the six participating practices, at least one general practitioner (GP) was recruited to complete pre- and post-study surveys and to take part in post-study interviews. A research nurse emailed GP participants the participant information sheet and survey link. Patients were invited to participate in qualitative interviews via an automated SMS sent by their practice, with instructions to contact the research team if interested.

### Informed consent

Practices provided organisational consent to participate. As Think-HF was designed to support clinicians in applying existing clinical guidance and did not alter care pathways, individual patient consent for CDST use was not required. Individual clinician consent was obtained prior to participation in surveys or interviews. Patient participants provided informed consent before completing post-consultation interviews.

### Baseline characteristics

Baseline practice surveys captured information on the composition of healthcare professionals involved in symptom assessment and HF diagnosis, and the availability of point-of-care natriuretic peptide (NP) testing. Baseline GP surveys collected information on years since qualification, clinical special interests, prior experience using decision support tools, HF-specific training, and confidence in applying HF guidance. Survey data were collected electronically via secure online links.

Practice-level anonymised data were also extracted on the number of registered patients, sociodemographic characteristics (age, sex, ethnicity), and prevalence of recorded HF diagnosis.

### Intervention: Think-HF CDST

Think-HF is an electronic clinical decision support tool remotely deployable within primary care IT systems using SystmOne. Once activated it uses coded data, prescriptions, test results and selected free-text terms to identify possible undiagnosed HF and highlight coding inconsistencies. For detailed definitions of all primary and secondary coding algorithms, see **Table S3**.

#### Triggering Mechanism

The primary trigger activates when a patient record is opened and the patient meets:

- ≥2 relevant comorbidity indicators, and
- ≥1 HF-suggestive symptom.

A prompt appears with options to: action, defer, cancel or pause (**Figure 1**).

**Figure 1:**
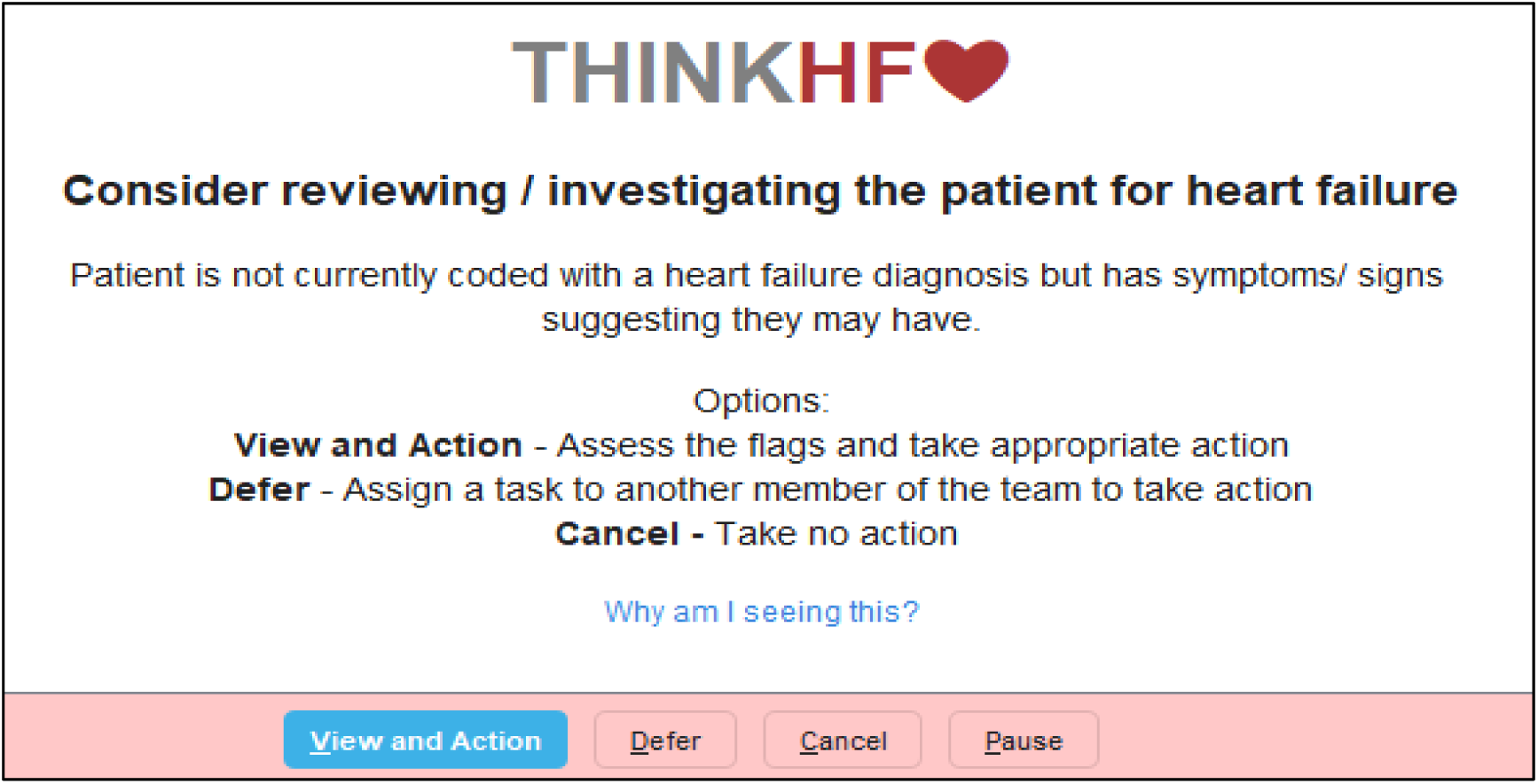
THINK HF alert.

#### Decision Support Template

If actioned, a structured template summarises HF-relevant clinical information (**Figure 2**) including:

- medical history,
- symptoms,
- existing or pending investigations, and
- prescribed medications.

**Figure 2:**
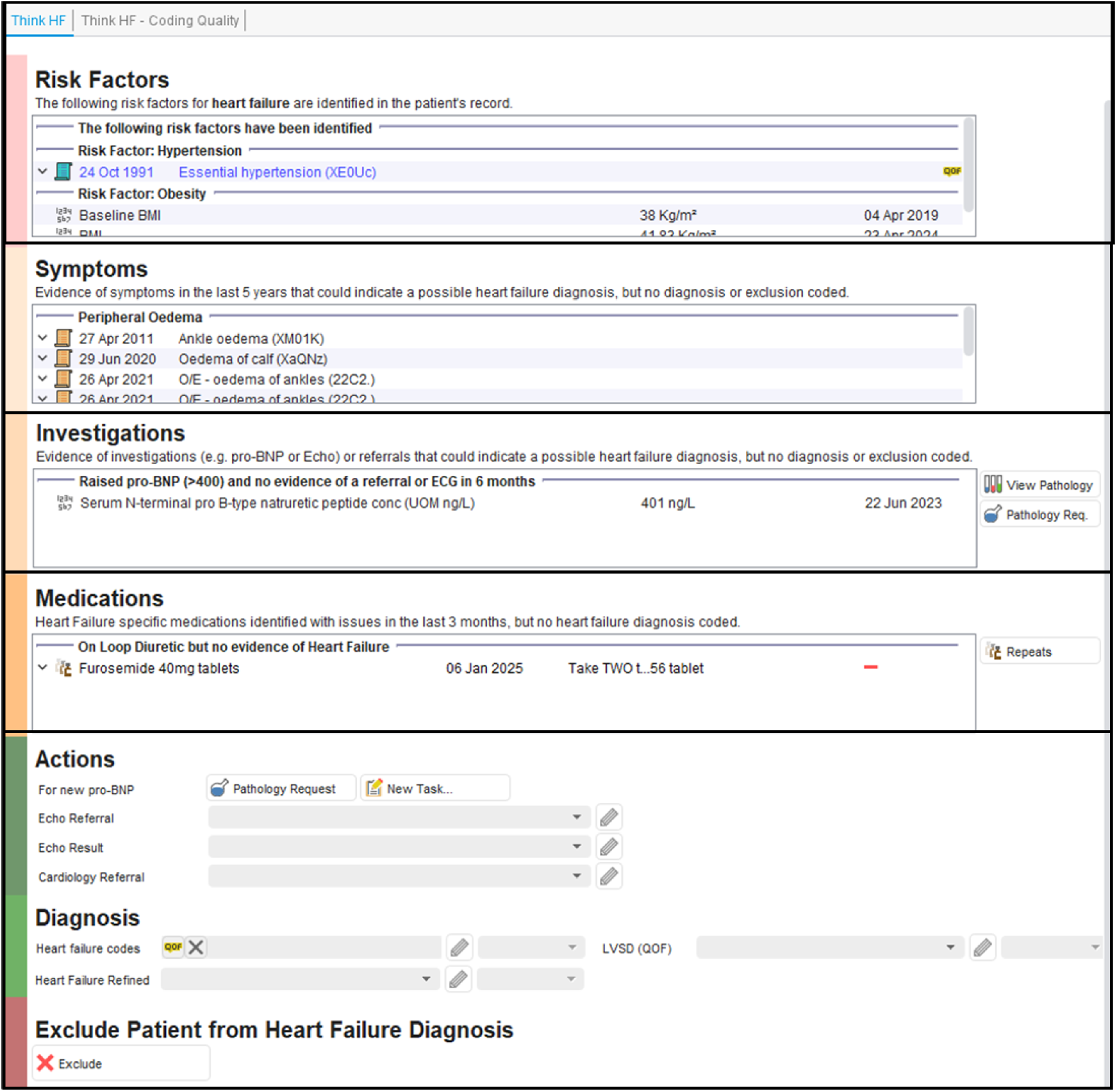
THINK HF template.

One-click actions enable the clinician to:

- order NP or echocardiography,
- refer to cardiology,
- apply HF diagnostic coding, or
- record exclusion of HF.

#### Secondary Algorithms

Additional algorithms identify patients with:

● unresolved investigations,
● inconsistent coding patterns, or
● medication profiles suggestive of HF.

These generate notification flags visible on the patient record homepage **(S1 Figure**). Selecting a flag opens a tailored template with suggested next actions. A template example for coding issues is provided in the supplementary information (**S2 Figure**).

### Training and Implementation Procedures

HCPs at each practice were invited to attend a 30-minute training session, delivered either in person or online. Training covered the purpose and evidence base for the tool, including findings from the epidemiological analysis, how and when alerts would appear, recommended actions, proactive use, and troubleshooting. A user guide and video were provided, and practices could contact the study team for support.

### Outcome Measures

Outcome measures focused on progression criteria for a future evaluation trial and on mechanisms influencing implementation, embedding, and integration of Think-HF into routine workflows, including barriers and enablers to modification for a definitive trial. Data sources included usage analytics, structured feedback forms, and interviews..

### Progression criteria

Progression criteria assessed feasibility across the core implementation dimensions of coverage and uptake, usability and acceptability, fidelity and implementation quality, and the potential for sustained engagement (**Table 1**). We also captured early indicators of effectiveness across the Think-HF algorithms, including changes in:

i. the number of patients added to the HF register;
ii. the proportion of HF patients with a refined diagnosis code;
iii. the number of patients with a raised natriuretic peptide test but no subsequent referral for echocardiography, echo result, or cardiology referral within the expected window (2 weeks prior to >6 months after)
iv. the number of patients prescribed loop diuretics without an HF diagnosis or exclusion code and with no history of a natriuretic peptide test.

**Table 1:** Progression criteria.

| PROGRESSION CRITERIA |  |  |  | STUDY DATA |
| --- | --- | --- | --- | --- |
| REACH |  |  |  |  |
| Number of practices recruited | 4 | <4 | None | Study data |
| Proportion of GPs willing to take part | >75% | 50-75% | <50% | Study data |
| Proportion of practices with > 100 patients identified by think-HF at baseline | 100% | 50-100% | <50% | Tool |
| How many patients with the main trigger consult per month per practice | >4 | 3-4 | <3 | Tool |
| EFFECTIVENESS |  |  |  |  |
| Data capture including clinical (MLTCs, appropriate diagnostic tests, medications) | >80% | 70-79% | >80% | Tool |
| ADOPTION |  |  |  |  |
| Acceptability of intervention: THINK-HF CDST perceived to be very or somewhat helpful for: |  |  |  |  |
| Patient alerts | >80% | 70-79% | <70% | Post survey 5 |
| Diagnostic prompts | >80% | 70-79% | <70% | Post survey 6 |
| HF diagnosis | >80% | 70-79% | <70% | Post survey 7 |
| Acceptability of training: THINK-HF training perceived to be very/somewhat helpful | >80% | 70-79% | <70% | Post survey 2 |
| IMPLEMENTATION |  |  |  |  |
| Correct installation | 100% | 75-99% | <75% | Study data |
| Proportion of eligible patients with indicators who trigger the alert during consultation | >90% | 70-89% | <70% | Study data |
| Prompts responded to by the GP | >40% | 30-40% | <30% | Tool & post survey 13 |
| MAINTANENCE |  |  |  |  |
| Proportion of practices retained in the study | All | >50% | <50% | Study data |
| GP, general practitioner; MLTCs, multiple long-term conditions; HF, heart failure; CDST, clinical decision support tool |  |  |  |  |

GP, general practitioner; MLTCs, multiple long-term conditions; HF, heart failure; CDST, clinical decision support tool

#### Process Evaluation

A mixed-methods process evaluation assessed acceptability and usability, feasibility and adoption patterns, fidelity and workflow integration, and perceived sustainability. Using the BEhavior and Acceptance fRamework (BEAR),(40) baseline and post-study clinician surveys (S3-S4) and semi-structured interview guides for clinicians and patients (S5-S6) were developed to identify, characterise, and explain mechanisms influencing implementation and integration of Think-HF. BEAR integrates the Theoretical Domains Framework(41) and the Unified Theory of Acceptance and Use of Technology(42) to bridge the gap between behavioural change and technology acceptance. All interviews were recorded, professionally transcribed and coded using NVivo software.

Post-intervention GP surveys collected data on perceived strengths and limitations of Think-HF and training, prompt response behaviour, and changes in confidence applying HF guidance. Process evaluation outcomes are summarised in Table 2.

**Table 2:** Process evaluation outcomes mapped to behavioural domains.

|  | Bear domains |  |  |  |  |  |  |  |  |  |  |  |  |  |  |  |  |  |  |  |  |  |
| --- | --- | --- | --- | --- | --- | --- | --- | --- | --- | --- | --- | --- | --- | --- | --- | --- | --- | --- | --- | --- | --- | --- |
|  | Knowledge | Skills, ability, and competence | Role and identity | Beliefs about capabilities | Beliefs about consequences | Attitudes | Contingencies | Intentions | Goals | Memory, attention, & decision | Environmental context and | Social influences | Emotions | Behavioural regulation | Intervention characteristics | Performance expectancy | Effort expectancy | Demographic characteristics | System quality | Agreement with the decision | Patient–health professional | Patient’ s preferences |
| REACH |  |  |  |  |  |  |  |  |  |  |  |  |  |  |  |  |  |  |  |  |  |  |
| Patient groups for whom the tool is perceived most useful |  |  |  |  |  | X |  |  |  |  |  |  |  |  |  | X |  |  |  |  |  |  |
| EFFECTIVENESS |  |  |  |  |  |  |  |  |  |  |  |  |  |  |  |  |  |  |  |  |  |  |
| Perceived effectiveness of the tool |  |  | X | X |  | X |  |  |  |  | X |  |  |  |  | X |  |  |  |  |  |  |
| ADOPTION |  |  |  |  |  |  |  |  |  |  |  |  |  |  |  |  |  |  |  |  |  |  |
| Prior experience with CDST and perceived usefulness | X | X |  |  |  | X |  |  |  |  |  |  |  |  |  | X | X |  |  |  |  |  |
| Prior experience with CDST for HF assessment/ management | X | X |  |  |  | X |  |  |  |  |  |  |  |  |  | X | X |  |  |  |  |  |
| Confidence in applying HF guidelines before and after study | X | X |  | X |  |  |  |  |  |  |  |  |  |  |  |  |  |  |  |  |  |  |
| Acceptability of intervention & perceived helpfulness in HF diagnosis. |  |  |  |  |  |  |  |  |  |  |  |  |  |  |  |  |  |  |  |  |  |  |
| Perceived importance of the tool for HF diagnosis and outcomes. | X |  | X | X |  | X | X | X | X | X | X |  |  |  | X |  |  |  |  |  |  |  |
| Strengths and limitations of THINK-HF CDST |  | X | X | X |  | X | X | X | X | X | X |  |  |  | X | X | X |  |  |  |  |  |
| Reasons for not following THINK HF prompts | X | X |  |  | X |  |  |  |  |  |  |  |  |  |  |  |  |  |  |  |  |  |
| Prior training in assessment and management of HF |  |  | X |  |  | X |  |  |  | X | X |  |  |  |  | X |  |  |  |  |  |  |
| Perceived quality of THINK HF training |  |  | X |  |  | X |  |  |  | X | X |  |  |  |  | X |  |  |  |  |  |  |
| Acceptability of THINK HF training |  | X |  | X |  | X |  |  |  |  |  |  |  |  |  |  |  |  |  |  |  |  |
| IMPLEMENTATION |  |  |  |  |  |  |  |  |  |  |  |  |  |  |  |  |  |  |  |  |  |  |
| Barriers and facilitators of implementation | X | X | X | X |  | X | X | X | X | X | X |  |  |  | X |  |  |  | X | X |  |  |
| MAINTANENCE |  |  |  |  |  |  |  |  |  |  |  |  |  |  |  |  |  |  |  |  |  |  |
| Considerations for longer-term embedding of the CDST across practices |  |  |  |  |  | X | X | X | X | X | X |  |  |  | X |  |  |  | X | X |  |  |
| CDST, clinical decision support tool; HF, heart failure. Bear, BEhavior and Acceptance fRamework. |  |  |  |  |  |  |  |  |  |  |  |  |  |  |  |  |  |  |  |  |  |  |

### Analysis

Interview transcripts underwent thematic analysis following Braun and Clarke’s six-step method.(43) Coding was initially inductive before being applied deductively to RE-AIM domains. Quantitative feasibility measures were summarised descriptively.

### Ethical review

The study was conducted in accordance with the principles outlined in the Declaration of Helsinki. Ethics approval was granted by the Health and Social Care Research Ethics Committee A (HSC REC A) [23/NI/0156].

#### Patient and public involvement

Patient and public involvement was integrated throughout the study. A patient advisory group comprising individuals with lived experience of HF contributed to study procedures, review of patient-facing materials, and refinement of interview content. Members were recruited via a national patient-led charity (Pumping Marvellous) with additional external recruitment to ensure diversity of perspectives. Their involvement ensured alignment with patient priorities and enhanced acceptability and relevance of study materials.

## Results

### Workflow Observations

Twenty-one consultations were observed across three GP practices (October-November 2024). Consultations lasted a median of 11.5 minutes (IQR 15). In all but three encounters, clinicians reviewed patient records in advance, typically examining the reason for attendance, past medical history, medications and recent test results. Of the 17 consultations in which patients reported symptoms (six involving breathlessness and one fluid retention) most symptoms were documented during the consultation using free text or templates rather than clinical codes; only three consultations resulted in a NP request. Symptom recording took place post-consultation in several cases and pre-consultation in one.

Clinical decision support alerts appeared in only four consultations and were limited to administrative reminders (e.g., flu vaccinations) rather than clinically actionable guidance. Half of all consultations involved interaction with the wider multidisciplinary team (MDT), largely via electronic tasking for tests or referrals. GPs highlighted limitations of existing alert systems, noting that some are intrusive, overly detailed or provide redundant information. Alerts perceived as most useful were those that surfaced concise, relevant clinical information such as recent test results. Opinions on the development of Think-HF was that it should surface key clinical data (including recent BNP results), recommend appropriate investigations (BNP and echocardiography), minimise mandatory data entry, and allow rapid dismissal to avoid workflow disruption.

### RAG-Based Refinement Exercise

Eleven HF clinical experts from national HF networks took part in the refinement of Think-HF algorithms including 3 senior primary care general practitioners, 5 senior cardiologists, 2 HF specialist nurses and 1 pharmacist. The RAG rating process refined the indicator set to ensure clinical relevance, specificity, and feasibility of implementation using routine primary care data.

- Symptoms: The clinical group recommended including specific HF-related symptoms (e.g., orthopnoea, paroxysmal nocturnal dyspnoea, peripheral oedema). Generic symptoms such as breathlessness were excluded due to low specificity, and weight gain was considered too inconsistently documented. Symptom searches were restricted to the prior five years and supplemented with free-text search capability.
- Comorbidities: Indicators were refined to emphasise cardiometabolic risk and disease clusters, with additions such as cardiomyopathies, chronic kidney disease and obstructive sleep apnoea. Arrhythmia indicators focused specifically on atrial fibrillation
- Physical Examination: Physical examination findings were excluded from the algorithm due to inconsistent recording, poor specificity, and limited structured coding.
- Diagnostic Tests: BNP testing was prioritised as a strong diagnostic indicator. Chest radiography was considered variably accessible and dependent on referral detail, while ECG and echocardiography were often non-specific or unavailable within structured primary care coded fields.
- Medications: Loop diuretics (furosemide, bumetanide, torasemide) were retained as a primary indicator, with additional HF-related medications (e.g., digoxin, sacubitril/valsartan, hydralazine/nitrate combinations, ivabradine, and SGLT2 inhibitors) flagged as potential supporting indicators.

The panel also noted important coding challenges, including missing HF phenotype codes and isolated diagnostic or functional codes (e.g., LV systolic dysfunction or diastolic dysfunction) without a formal HF diagnosis. These patterns were considered potential markers of missed or incomplete diagnostic pathways and were integrated into the logic of Think-HF.

### Feasibility results

#### Population

Six practices and 63 GPs participated in the feasibility study, with list sizes ranging from 8,350 to 25,128 patients. Four practices were located in more deprived neighbourhoods (IMD 1-2). Across all practices, the total registered population was 78,640: 38% were aged over 40 years, 47.5% were women, and the ethnic distribution was 59.4% White, 17.8% South Asian, 8.5% Black, 3% Chinese and 2.8% mixed ethnicities (Table 3). A total of 801 patients (1%) were on the HF register, of whom only 136 (17%) had a refined left ventricular phenotype code (43% HFrEF; 56% HFpEF).

**Table 3:** Practice and patient characteristics.

|  |  |  |  |  |  |
| --- | --- | --- | --- | --- | --- |
| **Practices** | **6** |  |  |  |  |
| IMD 1 (most deprived) | 1 |  |  |  |  |
| IMD 2 | 3 |  |  |  |  |
| IMD 3 | 2 |  |  |  |  |
| IMD 4 | - |  |  |  |  |
| IMD 5 (most affluent) | - |  |  |  |  |
| List size | Total 78,640; Median 11,371 (IQR 9,298); Range 8,350 to 25,128 |  |  |  |  |
| **Patients** | **78,640** |  |  |  |  |
| Male | 41,430 (52.7) |  |  |  |  |
| Female | 37,206 (47.3) |  |  |  |  |
|  |  | **Age** |  |  |  |
| 30-39 | 48,753 (62) | 60-69 | 5,913 (7.5) | >=90 | 352 (0.5) |
| 40-49 | 9,872 (12.6) | 70-79 | 3,883 (4.9) |  |  |
| 50-59 | 8,020 (10.2) | 80-89 | 1843 (2.3) |  |  |
|  |  | **Ethnicity** |  |  |  |
| White | 38,199 (59.4) | Chinese | 1,937 (3.0) | White/Asian | 687 (1.1) |
| Indian | 9740 (15.2) | Black Caribbean | 472 (0.7) | Mixed Other | 1,098 (1.7) |
| Pakistani | 1374 (2.1) | Black African | 3,820 (5.9) | Other | 2,212 (3.4) |
| Bangladeshi | 347 (0.5) | Black other | 1,058(1.7) |  |  |
| Asian other | 2,175 (3.4) |  |  |  |  |
| **HF register** | **801/78,640 (1.0)** |  |  |  |  |
| *Unrefined code* | 665/801 (83) |  |  |  |  |
| *Refined code* | 136/801 (17) |  |  |  |  |
| HFrEF | 58/136 (42.7) |  |  |  |  |
| HFmrEF | 2/136 (1.5) |  |  |  |  |
| HFpEF | 76/136 (55.9) |  |  |  |  |

All data presented as n (%) unless otherwise stated. IMD, index multiple deprivation; HFrEF, heart failure with reserved ejection fraction; HFpEF, heart failure with preserved ejection fraction;

Nine GPs participated in the process evaluation (6 men, 3 women; 1 White, 5 Asian and 3 identifying as ‘other’ or preferring not to say). Four had more than 10 years’ experience and five had less. Seven GPs had previous experience using CDSTs; all but one found them helpful for highlighting key information or providing rapid access to next steps. Most felt their training and clinical guidelines equipped them to assess and manage HFrEF, but only two felt similarly confident regarding HFpEF.

#### Baseline Think-HF indicators

At baseline, 876 patients (1.1%) met the main trigger criteria (≥2 comorbidities plus a recorded HF symptom). A further 6,838 patients (8.7%) had ≥2 risk factors without a recorded symptom and 2,805 (3.6%) met additional algorithm criteria (**Table 4**). The diagnostic consistency algorithms identified coding issues in 725 (83%) of the 801 patients already on the HF register and in 567 patients (0.7%) without a current HF diagnosis. Among those on the HF register with coding issues, the most common flag (62.6%) was a recorded left ventricular systolic dysfunction (LVSD) assessment without a subsequent refined HF phenotype code. In addition, 37% had no LVSD assessment recorded. Among the 567 patients with coding issues not on the HF register, the most frequent issues were the presence of diastolic dysfunction codes (62.1%) or HF administrative codes (33.5%; e.g., referrals or education) without any HF diagnosis or exclusion coding.

**Table 4:** Triggers existing in the GP databases at baseline.

| <u>Main trigger</u> | N (%) |
| --- | --- |
| ! THINK-HF Trigger (2 more risk factors + HF symptom) | 876/78640 (1.1) |
| Trigger for Free Text once symptom presents | 6838/78640 (8.7) |
| **<u>a. Coding Quality - Heart Failure patients (n=801) with coding quality issues</u>** | **725/801 (90.5)** |
| b. HF QOF Register but no QOF LVSD assessment | 265/725 (36.6) |
| c. HF QOF Register & LVSD Assessment but no HF Refined code | 454/725 (62.6) |
| d. HF diagnosed but has a possible normal subsequent echocardiogram - Consider new HF code, HF Refined code or add resolved/exclude | 25/725 (3.4) |
| **<u>e. Patients that are very likely HF but not coded</u>** | **567/78640 (0.7)** |
| f. LVSD codes but no HF diagnosis or exclusion codes | 85/567 (15.0) |
| g. Diastolic Dysfunction codes but no HF diagnosis or exclusion codes | 352/567 (62.1) |
| h. HF Proxy codes but no HF diagnosis or exclusion codes | 190/567 (33.5) |
| **<u>i. Investigation based HF case finding</u>** | **960/78640 (1.2)** |
| j. Raised pro-BNP, no echocardiogram or result and no referral to cardiology between 2 weeks before to 6 months after. | 332/960 (34.6) |
| k. Raised pro-BNP with latest echocardiogram >6 months prior and no result | 0 |
| l. No pro-BNP after symptoms code and has risk factors | 660/960 (68.8) |
| **<u>m. Medication based HF case finding</u>** | **553/78640 (0.7)** |
| n. Entresto / Sacubitril and no HF diagnosis or exclusion codes | 17/553 (3.1) |
| n. Hydralazine in combination with Nitrate but no HF diagnosis or exclusion codes | 0 |
| o. Ivabradine but no HF diagnosis or exclusion codes | 20/553 (3.6) |
| p. SGLT2i but no Diabetes diagnosis or HF diagnosis or exclusion codes | 43/553 (7.8) |
| q. On Digoxin and no AF diagnosis or HF diagnosis or exclusion codes | 2/553 (0.4) |
| w. 1. Loop Diuretics but no HF diagnosis or exclusion codes | 490/553 (88.6) |
| w. 2. Loop Diuretics but no HF diagnosis or exclusion code AND abnormal pro-BNP | 87/553 (15.7) |
| w. 3. Loop Diuretics but no HF diagnosis or exclusion code AND no history of pro-BNP | 210/553 (38) |
| All data presented as n (%) unless otherwise stated. HF, heart failure; QOF, quality outcomes framework; LVSD, left ventricular systolic dysfunction; SGLT2i, Sodium-Glucose Cotransporter-2 Inhibitors; AF, atrial fibrillation; BNP, B-type Natriuretic Peptide |  |

Unresolved investigations were identified in 960 patients (1.2%). Of these, 34.6% had a raised NP without a subsequent referral for echocardiography, an echo result, or referral to cardiology within the expected window (2 weeks before to 6 months after). A further 69% had ≥2 comorbidities plus a HF symptom but no pro-BNP test. Medication-based issues were present in 553 patients (0.7%), predominantly driven by loop diuretic prescribing without a HF diagnosis (88.6%), and by patients prescribed a loop diuretic without a pro-BNP test (38%) and no HF diagnosis or exclusion code.

#### Progression criteria

*Reach*: Of the 15 GP practices approached, six were recruited (40% recruitment rate), exceeding the target of four practices. Of the 13 GPs invited to participate in the process evaluation, nine (69%) agreed. All participating practices had more than 100 patients identified by Think-HF at baseline (range: 259-1,556).

During the 4-month trial period, 929 patients across participating practices met the main trigger criteria, of whom 299 (32.2%) triggered an alert, indicating that the clinical record had been opened. Of these 299 triggers, 224 (75%) occurred on the same day as a clinical consultation. On average, nine patients per month per practice triggered the main alert during a consultation, exceeding the prespecified target of four. However, feedback from the first two practices indicated that the alert was activating too frequently. In response, the main trigger was refined to activate only for patients with a newly recorded comorbidity or symptom, while patients with historical indicators were assigned a non-interruptive flag within their record. Following this modification, the average number of consultation-based alerts fell from 20 to 4 per month per practice, aligning with expected levels and improving feasibility for routine clinical use.

#### Effectiveness

All data were captured accurately by the CDST. Across early effectiveness indicators, Think-HF demonstrated positive shifts in diagnostic coding and investigation pathways (**Table 5**). Over the 4-month trial, 31 patients were added to the HF register, representing a 4.2% relative increase. The largest increase (+10%) occurred in the first two practices, where baseline HF prevalence was lower (0.67%) compared with the remaining four practices (1.28%). Among patients diagnosed with HF, the number with a refined HF diagnosis code increased from 136 to 159, corresponding to a 14% relative increase.

**Table 5:** Early effectiveness indicators.

|  | All practices |  |  | First two practices |  |  | Second four practices |  |  |
| --- | --- | --- | --- | --- | --- | --- | --- | --- | --- |
|  | Baseline | Post-trial | Relative change | Baseline | Post-trial | Relative change | Baseline | Post-trial | Relative change |
| Population | 78640 | 78398 |  | 33602 | 33680 |  | 45038 | 44718 |  |
| Number of patients on HF QOF Register | 801/7864<br>0 (1.02) | 832/7839<br>8 (1.06) | +4.19% | 224/3360<br>2 (0.67) | 247/3368<br>0 (0.73) | +10.01<br>% | 577/4503<br>8 (1.28) | 585/4471<br>8 (1.31) | +2.1% |
| Number of patients with HF phenotype code | 136/810<br>(16.79) | 159/832<br>(19.11) | +13.82<br>% | 41/224<br>(18.30) | 50/247<br>(22.32) | +21.95<br>% | 95/577<br>(16.46) | 109/585<br>(18.63) | +13.2% |
| Raised pro-BNP but no referral to echo, echo result, or referral to cardiology between 2 weeks before to 6 months after. | 332/7864<br>0 (0.42) | 309/7839<br>8 (0.39) | -6.64% | 91/33602<br>(0.27) | 96/33680<br>(0.29) | +5.25% | 241/4503<br>8 (0.54) | 213/4471<br>8 (0.48) | -10.99% |
| Loop Diuretics but no HF diagnosis or exclusion code AND no history of pro-BNP | 210/7864<br>0 (0.27) | 181/7839<br>8 (0.23) | -13.54% | 42/33602<br>(0.12) | 23/33680<br>(0.07) | -45.36% | 168/4503<br>8 (0.37) | 158/4471<br>8 (0.35) | -4.8% |

All data presented as n (%) unless otherwise stated. HF, heart failure; QOF, quality outcomes framework; BNP, B-type Natriuretic Peptide

The number of patients with an elevated pro-BNP but no subsequent echocardiography referral, recorded echocardiogram, or cardiology referral within the expected timeframe (<2 weeks to >6 months) decreased by 23 patients (7% relative reduction). Similarly, the number of patients prescribed a loop diuretic without an HF diagnosis or exclusion code and with no prior pro-BNP testing fell by 29 patients, representing a 14% relative reduction. The most substantial improvement was observed during the first implementation iteration, when pharmacist-led support addressing medication-related issues in one participating practice contributed to a 45% reduction in this category.

#### Adoption

All GPs reported that the Think-HF training, alerts, test-prompting and diagnostic support were somewhat or very helpful.

#### Implementation and Maintenance

The tool was correctly installed in all practices and triggered for all eligible patients during consultations. Of the 299 alerts generated, GPs recorded a response (actioned or deferred) for 131 (44%). The response rate increased from 41.7% in the first iteration to 50% in the second, following optimisation to reduce alert frequency. Overall, two GPs reported following the prompts sometimes, three a few times, and two most of the time. All practices were retained in the study.

#### Process evaluation

*GPs* **(See S1 Table for related quotes)**

#### Reach and Effectiveness

Clinicians reported that Think-HF reached the right patients, particularly those with multiple long-term conditions (MLTCs), overlapping respiratory or liver conditions or obesity, where HF can easily be overlooked. Prompts helped surface cases where symptoms such as breathlessness or oedema had not previously triggered HF assessment, supporting identification of undiagnosed patients. Early diagnosis was viewed as essential for preventing deterioration, avoiding emergency admissions and enabling timely optimisation of therapy. Think-HF contributed to this by providing clear, well-timed cues that redirected attention to HF during busy consultations, highlighting missed investigations and encouraging more systematic work-up, coding and referral. Several clinicians described instances where the tool directly changed clinical decisions or prompted earlier investigation.

#### Adoption and Implementation

Adoption was broadly positive, with prompts described as relevant, specific and easy to use. However, engagement varied depending on workload and alert frequency. Time pressure in 10-minute consultations meant clinicians often deferred prompts for later review, using task-sharing approaches such as sending cases to a nominated colleague or batching reviews outside clinic time. Some also noted that routine long-term condition reviews conducted by practice nurses created opportunities for symptom recognition and escalation, indicating potential for wider MDT involvement. Ongoing support, including refresher training, reminders and help running searches, was viewed as important for maintaining consistency.

While overall integration was strong, clinicians noted alert fatigue and the visible ‘footprint’ left when dismissing prompts as areas for refinement.

#### Maintenance

Clinicians were optimistic about sustaining Think-HF in routine practice. The tool increased awareness of HF and encouraged more proactive diagnostic thinking. Continued optimisation and training, whole-team engagement and reliable access to investigations (particularly BNP and echocardiography) were identified as important for long-term maintenance. Overall, Think-HF was considered feasible to embed at scale and likely to improve early recognition of HF.

#### Process evaluation

*Patients* **(See S2 Table for related quotes)**

Patient interviews explored experiences of seeking assessment, navigating primary care processes and receiving explanations around symptoms and diagnoses. Several themes are directly relevant to the optimisation of Think-HF. Access mechanisms such as early morning telephone queues, online forms and reception triage strongly influenced whether symptoms were assessed in a timely way, with digital barriers and mobility issues often delaying review. Continuity of care was highly valued but frequently inconsistent; many patients described repeating their history to multiple clinicians, while trusted relationships with a known GP or a nurse facilitated clearer plans and earlier escalation when needed.

Communication gaps were common, with patients often unaware of test results, unclear about next steps or needing to ‘chase’ appointments. Delayed or misattributed symptoms were frequently described including breathlessness, palpitations or fatigue which were often initially attributed to anxiety, ageing or respiratory causes.

## Discussion

This mixed-methods feasibility study demonstrates that Think-HF, a novel electronic decision-support tool designed to identify possible undiagnosed HF in primary care, is both usable and acceptable and has the potential to strengthen early diagnostic pathways. Across six practices serving socioeconomically and ethnically diverse populations, Think-HF was deployed successfully, reached clinically relevant patient groups, and prompted earlier consideration of HF in presentations where symptoms are often misattributed or overlooked. Early indicators of effectiveness, including refined HF coding, fewer unresolved investigations and improved follow-up of medication-based signals, suggest that Think-HF can improve key steps in the diagnostic pathway. These findings align with the NHS 10-Year Plan and its three shifts, supporting proactive prevention, earlier diagnosis, digital enablement and care closer to home. Combined clinician and patient insights support progression to larger-scale evaluation.

Three overarching findings emerged. First, Think-HF reliably reached patients most likely to benefit: those with MLTCs, overlapping respiratory, renal or hepatic conditions and obesity that complicate symptom interpretation. Clinicians consistently described the tool as issuing relevant prompts that surfaced HF as a diagnostic consideration at appropriate times, mirroring epidemiological evidence that these groups are disproportionately under-diagnosed.(34) Second, Think-HF influenced clinical behaviour, aligning with evidence that integrated, actionable decision support improves guideline-concordant care.(27,44) Prompts acted as cognitive ‘nudges’ known to improve guideline adherence.(45,46) During time-pressured consultations, this can help refocus attention on HF where symptoms may otherwise be overshadowed. Clinicians described multiple instances where Think-HF altered decisions, prompting BNP requests, reviewing investigations, revisiting differential diagnoses or correcting coding inconsistencies. Third, implementation was feasible. Although high frequency of alerts can lead to ‘alert fatigue’,(47) once the tool had been refined, the tool aligned well with routine workflows. Practices adopted strategies for managing alerts, including task-sharing, batching cases and drawing on nurse-led long-term condition reviews. These adaptations reflect primary care workload and highlight opportunities for broader MDT involvement.

The study reinforces the challenge of under-recognition of HF in primary care, particularly for patients with MLTCs. Think-HF addressed known barriers: low specificity of early symptoms such as breathlessness,(48,49) inconsistent coding,(50,51) and delays in the follow-up of abnormal investigations.(52) By providing timely prompts and a structured template, the tool helped clinicians navigate diagnostic uncertainty, improve coding accuracy, encourage earlier initiation of guideline-recommended investigations, and flag unresolved investigations. For patients, these improvements matter. Interviews highlighted diagnostic delays, frequent misattribution of symptoms, and variable communication at key points in the pathway, reflecting prior reports.(8,53,54) Many relied on repeated presentations or self-advocacy before diagnosis. Tools such as Think-HF can help reduce these disparities by ensuring early HF signals are consistently recognised and acted upon, regardless of clinician or workload pressures.

The process evaluation highlights considerations for wider implementation. Workload pressures influenced use, with many clinicians reviewing prompts outside of consultations. This underscores the importance of embedding tools within whole-team workflows,(52,53) enabling shared responsibility for reviewing prompts, arranging investigations and ensuring follow-up.

Clear opportunities for MDT involvement in Think-HF were identified. Practice nurses, pharmacists, care coordinators and administrative staff already contribute to long-term condition reviews, medication management and pathway navigation. Think-HF’s structured template aligns with these roles, and its task-sharing functionality supports distributed diagnostic safety-netting. In one practice, pharmacist-led follow-up reduced loop-diuretic prescribing without natriuretic peptide testing by 45%. Broader use across the MDT could enhance timeliness, consistency and sustainability. Similar models, such as PINCER,(26) which utilised pharmacist-led CDST support reduced hazardous prescribing by up to 49%, demonstrating the value of shared responsibility in clinical risk reduction.

Clinicians emphasised that ongoing education and support are essential. Although initial training was well received, sustained use will require regular refreshers, micro-learning resources, practice-wide updates and shared learning.(55,56) HF guidance evolves and staff turnover is inevitable, making continuous educational support key. Practices also highlighted the need for support in running searches, resolving coding anomalies and integrating Think-HF into quality-improvement activity. The potential for a local champion role emerged informally in several practices. Champions can support colleagues and coordinate education, although evidence for their impact remains mixed.(57,58) In Think-HF, champions should form part of a broader implementation package including whole-team engagement, MDT task-sharing and structured education.

System dependencies also remain important. Even when Think-HF prompts were acted upon, delays in referrals and complexities in echocardiography reports could slow diagnostic progression.(9,50) Improving diagnostic capacity, standardising reporting and ensuring timely review will be critical for maximal impact.(59) Patient perspectives underscore the importance of embedding digital tools in accessible, person-centred pathways.(60) Ensuring that MDT members, not only GPs, can act on Think-HF outputs provides multiple entry points for assessment and reduces reliance on repeated patient advocacy.

The study has several strengths. Think-HF was co-designed with clinicians, patients and informatics experts, drawing on extensive epidemiological evidence and consensus methods. Feasibility testing was conducted across diverse practices (two thirds in the most deprived areas and 40% non-White ethnicity), enhancing external validity. The mixed-methods approach captured complementary insights into usability, behavioural impact, acceptability and contextual influences, and the RE-AIM framework ensured structured assessment of implementation. However, the study also has limitations. The four-month follow-up period limited assessment of effectiveness or long-term diagnostic outcomes. Participating practices may have been more research-active than average, potentially inflating adoption. Further alert optimisation may be needed to balance sensitivity and specificity across broader populations. Think-HF was tested only in SystmOne practices, and adaptation may be required for use in other systems. Patient interviews, while rich, may not represent all groups at risk of delayed HF recognition. Finally, early effectiveness measures were descriptive; a controlled evaluation will be required to determine clinical impact on diagnostic rates, guideline optimisation and outcomes.

Think-HF is feasible, acceptable and well aligned with primary care workflows. By prompting timely consideration of HF, supporting structured assessment and improving coding and follow-up, it has clear potential to strengthen early diagnostic pathways. Expanding Think-HF to the wider MDT and providing structured educational support will be essential for sustainable adoption, providing a strong foundation for larger-scale evaluation.

## Data sharing

The data underlying this article cannot be shared publicly due to the privacy of individuals that participated in the study. The anonymised data, codelists and study protocol will be shared on reasonable request to the corresponding author.

## Author contributions

All authors had full access to all data in the study and were responsible for the decision to submit the manuscript and both CAL and KB accessed and verified the data. The senior author (CAL) affirms that the manuscript is an honest, accurate, and transparent account of the study being reported; that no important aspects of the study have been omitted; and that any discrepancies from the study as originally planned have been explained. KB: design, analysis, interpretation of data, co-writing original draft and revised drafts. CAL: conception, design, acquisition, analysis, interpretation of data, co-writing original draft and revised drafts. GPM, KK: conception, design, interpretation, resources, critical review. CD, SP: conception, design, interpretation, critical review. JGFC: conception, interpretation, critical review. CJT, LB, IS: interpretation, critical review.

## Acknowledgements

CL, KK and GPM are supported by the NIHR Leicester Biomedical Research Centre (BRC) and the British Heart Foundation Research Excellence Award (RE/24/130031). KK and CL are also supported by the National Institute for Health Research (NIHR) Applied Research Collaboration East Midlands (ARC EM) and KK by the NIHR Global Research Centre for Multiple Long-Term Conditions, NIHR Cross NIHR Collaboration for Multiple Long-Term Conditions. LB is an NIHR Senior Research Leader for Nursing and Midwifery. The views expressed in this publication are those of the authors and not necessarily those of the NIHR, NHS or the UK Department of Health and Social Care.

## References

1. Organisation for Economic Co-operation and Development. Health at a glance: Europe 2018. Paris: OECD/EU; 2018.

2. Alex Bottle, Dani Kim, Paul Aylin, Martin R Cowie, Azeem Majeed, Benedict Hayhoe. Routes to diagnosis of heart failure: observational study using linked data in England. Heart 2018;104(7):600–605.

3. Cuthbert JJ, Soyiri I, Lomax SJ, Turgoose J, Fuat A, Cohen J, et al. Outcomes in patients treated with loop diuretics without a diagnosis of heart failure: a retrospective cohort study. Heart 2024 May 23;110(12):854–862.

4. Friday JM, Cleland JGF, Pellicori P, Wolters MK, McMurray JJV, Jhund PS, et al. Loop diuretic therapy with or without heart failure: impact on prognosis. Eur Heart J 2024;45(37):3837–3849.

5. Lawson CA, Zaccardi F, Squire I, Ling S, Davies MJ, Lam CSP, et al. 20-year trends in cause-specific heart failure outcomes by sex, socioeconomic status, and place of diagnosis: a population-based study. The Lancet Public Health 2019;4(8):e406–e420.

6. Bachtiger P, Kelshiker MA, Petri CF, Gandhi M, Shah M, Kamalati T, et al. Survival and health economic outcomes in heart failure diagnosed at hospital admission versus community settings: a propensity-matched analysis. BMJ Health Care Inform 2023;30(1):e100718.

7. Taylor CJ, Ordóñez-Mena JM, Roalfe AK, Lay-Flurrie S, Jones NR, Marshall T, et al. Trends in survival after a diagnosis of heart failure in the United Kingdom 2000-2017: population based cohort study. BMJ 2019;364:l223.

8. Santos GC, Liljeroos M, Dwyer AA, Jaques C, Girard J, Strömberg A, et al. Symptom perception in heart failure: a scoping review on definition, factors and instruments. Eur J Cardiovasc Nurs 2020 Feb;19(2):100–117.

9. Kwok CS, Burke H, McDermott S, Welsh V, Barker D, Patwala A, et al. Missed Opportunities in the Diagnosis of Heart Failure: Evaluation of Pathways to Determine Sources of Delay to Specialist Evaluation. Curr Heart Fail Rep 2022 Aug;19(4):247–253.

10. Borlaug BA. Evaluation and management of heart failure with preserved ejection fraction. Nat Rev Cardiol 2020 Sep;17(9):559–573.

11. Gupta M, Bell A, Padarath M, Ngui D, Ezekowitz J. Physician Perspectives on the Diagnosis and Management of Heart Failure With Preserved Ejection Fraction. CJC Open 2020 Nov 16;3(3):361–366.

12. Hancock HC, Close H, Fuat A, Murphy JJ, Hungin APS, Mason JM. Barriers to accurate diagnosis and effective management of heart failure have not changed in the past 10 years: a qualitative study and national survey. BMJ Open 2014 Apr 1;4(3):e003866–003866.

13. Bozkurt B, Coats AJ, Tsutsui H, Abdelhamid M, Adamopoulos S, Albert N, et al. Universal Definition and Classification of Heart Failure: A Report of the Heart Failure Society of America, Heart Failure Association of the European Society of Cardiology, Japanese Heart Failure Society and Writing Committee of the Universal Definition of Heart Failure. J Card Fail 2021 Mar 1.

14. National Guideline Centre (. Chronic Heart Failure in Adults: Diagnosis and Management. London: National Institute for Health and Care Excellence (NICE). 2018 Sep.

15. McDonagh TA, Metra M, Adamo M, Gardner RS, Baumbach A, Böhm M, et al. 2021 ESC Guidelines for the diagnosis and treatment of acute and chronic heart failure: Developed by the Task Force for the diagnosis and treatment of acute and chronic heart failure of the European Society of Cardiology (ESC) With the special contribution of the Heart Failure Association (HFA) of the ESC. Eur Heart J 2021;42(36):3599–3726.

16. Sandhu AT, Tisdale RL, Rodriguez F, Stafford RS, Maron DJ, Hernandez-Boussard T, et al. Disparity in the Setting of Incident Heart Failure Diagnosis. Circ Heart Fail 2021 Aug;14(8):e008538.

17. Hayhoe B, Kim D, Aylin PP, Majeed FA, Cowie MR, Bottle A. Adherence to guidelines in management of symptoms suggestive of heart failure in primary care. Heart 2019 May;105(9):678–685.

18. Conrad N, Judge A, Canoy D, Tran J, O’Donnell J, Nazarzadeh M, et al. Diagnostic tests, drug prescriptions, and follow-up patterns after incident heart failure: A cohort study of 93,000 UK patients. PLOS Medicine 2019;16(5):e1002805.

19. Lawson CA, Ali MR, McCann GP, Squire I, Zaccardi F, Rashid M, et al. Health inequalities and trends in heart failure diagnosis in primary care in England, 2000-21: a national retrospective cohort data-linkage study. Lancet Prim Care 2025 Dec;1(6):None.

20. Murphy JJ, Chakraborty RR, Fuat A, Davies MK, Cleland JGF. Diagnosis and management of patients with heart failure in England. Clin Med (Lond) 2008 Jun;8(3):264–266.

21. De Clercq L, Himmelreich JCL, Harskamp RE. Quality of heart failure registration in primary care: observations from 1 million electronic health records in the Amsterdam Metropolitan Area. Diagnosis (Berl) 2024 May 14;11(4):380–388.

22. Klein S, Mukhopadhyay A, Hamo CE, Li X, Adhikari S, Blecker S. Accuracy of Electronic Health Record-Based Definitions for Patients with Heart Failure. Am J Med 2025;138(11):1557–1560.e1.

23. Balas EA, Weingarten S, Garb CT, Blumenthal D, Boren SA, Brown GD. Improving preventive care by prompting physicians. Arch Intern Med 2000 Feb 14;160(3):301–308.

24. Garg AX, Adhikari NKJ, McDonald H, Rosas-Arellano MP, Devereaux PJ, Beyene J, et al. Effects of computerized clinical decision support systems on practitioner performance and patient outcomes: a systematic review. JAMA 2005 Mar 9;293(10):1223–1238.

25. Schedlbauer A, Prasad V, Mulvaney C, Phansalkar S, Stanton W, Bates DW, et al. What evidence supports the use of computerized alerts and prompts to improve clinicians’ prescribing behavior? J Am Med Inform Assoc 2009;16(4):531–538.

26. Avery AJ, Rodgers S, Cantrill JA, Armstrong S, Cresswell K, Eden M, et al. A pharmacist-led information technology intervention for medication errors (PINCER): a multicentre, cluster randomised, controlled trial and cost-effectiveness analysis. The Lancet 2012;379(9823):1310–1319.

27. Shiffman RN, Liaw Y, Brandt CA, Corb GJ. Computer-based guideline implementation systems: a systematic review of functionality and effectiveness. J Am Med Inform Assoc 1999;6(2):104–114.

28. Kawamoto K, Houlihan CA, Balas EA, Lobach DF. Improving clinical practice using clinical decision support systems: a systematic review of trials to identify features critical to success. BMJ 2005 Apr 2;330(7494):765.

29. Ghazi L, Yamamoto Y, Riello RJ, Coronel-Moreno C, Martin M, O’Connor KD, et al. Electronic Alerts to Improve Heart Failure Therapy in Outpatient Practice: A Cluster Randomized Trial. J Am Coll Cardiol 2022 Jun 7;79(22):2203–2213.

30. Alderwick H. Government’s 10 year plan for the NHS in England. BMJ 2025 Jul 4;390:r1396.

31. Smeets M, De Witte P, Peters S, Aertgeerts B, Janssens S, Vaes B. Think-aloud study about the diagnosis of chronic heart failure in Belgian general practice. BMJ Open 2019 Mar 20;9(3):e025922–025922.

32. McEntee ML, Cuomo LR, Dennison CR. Patient-, provider-, and system-level barriers to heart failure care. J Cardiovasc Nurs 2009;24(4):290–298.

33. Lawson CA, Zaccardi F, Squire I, Okhai H, Davies M, Huang W, et al. Risk Factors for Heart Failure: 20-Year Population-Based Trends by Sex, Socioeconomic Status, and Ethnicity. Circ Heart Fail 2020 Feb;13(2):e006472.

34. Wong CW, Tafuro J, Azam Z, Satchithananda D, Duckett S, Barker D, et al. Misdiagnosis of Heart Failure: A Systematic Review of the Literature. J Card Fail 2021;27(9):925–933.

35. Beishon L, Jayasinghe R, Roshan-Zamir S, Barton C. Reducing heart failure deaths by 25% in 25 years: the ‘25in25’ heart failure summit. Br J Cardiol 2024 Jun 11;31(2):022.

36. Roche, Pumping Marvellous. Heart failure: The hidden costs of late diagnosis A report to identify the opportunities for earlier diagnosis. 2020 August.

37. Barber K, Bernhardt L, McCann GP, Squire I, Miller CA, Deaton C, et al. Developing core indicators for identifying people at risk of delayed heart failure diagnosis. BMC Prim Care 2025 Oct 16;26(1):316–4.

38. Glasgow RE, Harden SM, Gaglio B, Rabin B, Smith ML, Porter GC, et al. RE-AIM Planning and Evaluation Framework: Adapting to New Science and Practice With a 20-Year Review. Front Public Health 2019 Mar 29;7:64.

39. Moore GF, Audrey S, Barker M, Bond L, Bonell C, Hardeman W, et al. Process evaluation of complex interventions: Medical Research Council guidance. BMJ 2015 Mar 19;350:h1258.

40. Camacho J, Zanoletti-Mannello M, Landis-Lewis Z, Kane-Gill SL, Boyce RD. A Conceptual Framework to Study the Implementation of Clinical Decision Support Systems (BEAR): Literature Review and Concept Mapping. J Med Internet Res 2020 Aug 6;22(8):e18388.

41. Michie S, Johnston M, Abraham C, Lawton R, Parker D, Walker A. Making psychological theory useful for implementing evidence based practice: a consensus approach. Qual Saf Health Care 2005;14(1):26–33.

42. Venkatesh V, Morris MG, Davis GB, Davis F &. User Acceptance of Information Technology: Toward a Unified View. MIS Quarterly 2003 September 1;27(3):425–478.

43. Braun V, Clarke V. Using thematic analysis in psychology. Qualitative Research in Psychology 2006;3(2):77–101.

44. Kwan JL, Lo L, Ferguson J, Goldberg H, Diaz-Martinez JP, Tomlinson G, et al. Computerised clinical decision support systems and absolute improvements in care: meta-analysis of controlled clinical trials. BMJ 2020;370:m3216.

45. Nwafor O, Singh R, Collier C, DeLeon D, Osborne J, DeYoung J. Effectiveness of nudges as a tool to promote adherence to guidelines in healthcare and their organizational implications: A systematic review. Soc Sci Med 2021 Oct;286:114321.

46. Yoong SL, Hall A, Stacey F, Grady A, Sutherland R, Wyse R, et al. Nudge strategies to improve healthcare providers’ implementation of evidence-based guidelines, policies and practices: a systematic review of trials included within Cochrane systematic reviews. Implement Sci 2020 Jul 1;15(1):50–0.

47. Gani I, Litchfield I, Shukla D, Delanerolle G, Cockburn N, Pathmanathan A. Understanding “Alert Fatigue” in Primary Care: Qualitative Systematic Review of General Practitioners Attitudes and Experiences of Clinical Alerts, Prompts, and Reminders. J Med Internet Res 2025 Feb 7;27:e62763.

48. Mant J, Doust J, Roalfe A, Barton P, Cowie MR, Glasziou P, et al. Systematic review and individual patient data meta-analysis of diagnosis of heart failure, with modelling of implications of different diagnostic strategies in primary care. Health Technol Assess 2009 Jul;13(32):1–207, iii.

49. van Riet EES, Hoes AW, Limburg A, Landman MAJ, van der Hoeven H, Rutten FH. Prevalence of unrecognized heart failure in older persons with shortness of breath on exertion. Eur J Heart Fail 2014;16(7):772–777.

50. Crundall A, Crawshaw-Ralli M, Fuat A, Oberoi K, Crossan J, Jones S, et al. Lessons learnt from HF coding in primary care. What might best practice look like? Br J Cardiol 2024 Sep 6;31(3):037.

51. Cuthbert JJ, Gopal J, Crundall-Goode A, Clark AL. Are there patients missing from community heart failure registers? An audit of clinical practice. Eur J Prev Cardiol 2019 Feb;26(3):291–298.

52. Litchfield I, Bentham L, Lilford R, McManus RJ, Hill A, Greenfield S. Test result communication in primary care: a survey of current practice. BMJ Qual Saf 2015;24(11):691–699.

53. Park J, Seo K, Lee JE, Kim K, Ahn J. Communication needs regarding heart failure trajectory and palliative care between patients and healthcare providers: A cross-sectional study. PLoS One 2025 Jan 13;20(1):e0317417.

54. Taylor CJ, Hobbs FDR, Marshall T, Leyva-Leon F, Gale N. From breathless to failure: symptom onset and diagnostic meaning in patients with heart failure—a qualitative study. BMJ Open 2017;7(3):e013648.

55. De Gagne JC, Park HK, Hall K, Woodward A, Yamane S, Kim SS. Microlearning in Health Professions Education: Scoping Review. JMIR Med Educ 2019 Jul 23;5(2):e13997.

56. Ingram Nissen T, Edelman EA, Steinmark L, Logan K, Reed EK. Microlearning: Evidence-based education that is effective for busy professionals and short attention spans. J Genet Couns 2024 Feb;33(1):232–237.

57. Lüchinger R, Audétat M, Blondon K, Junod Perron N. Implementing collaborative practices in healthcare settings using champions: a scoping review. Implement Sci 2025 Nov 4;20(1):48–2.

58. Santos WJ, Graham ID, Lalonde M, Demery Varin M, Squires JE. The effectiveness of champions in implementing innovations in health care: a systematic review. Implement Sci Commun 2022 Jul 22;3(1):80–0.

59. Paton MF, Barton C, Baruah R, Hartshorne-Evans N, Jenkins GH, Potter A, et al. Echocardiography reporting in heart failure with preserved ejection fraction: Delphi consensus study. Open Heart 2025 Mar 28;12(1):e003063. doi: 10.1136/openhrt-003063.

60. Granström E, Wannheden C, Brommels M, Hvitfeldt H, Nyström ME. Digital tools as promoters for person-centered care practices in chronic care? Healthcare professionals’ experiences from rheumatology care. BMC Health Serv Res 2020 Dec 1;20(1):1108–5.

