## Supplementary information for "Think-HF: Development, feasibility, and usability of an electronic health record integrated tool for identifying missed diagnoses of heart failure in primary care"

### Supplementary File

#### THINK-HF: Development, feasibility, and usability study of an intelligent electronic health record integrated tool for early heart failure diagnosis in primary care

*Barber K, Deaton C, McCann GP, Bernhardt L, Prinjha S, Alaei Kalajahi R, Ali MR, Squire I, Taylor CJ, Cleland JGF, Khunti K, Lawson C.*

### Supplementary methods

#### S1 Method: Clinical observation of GP consultations template

| | |
|---|---|
| Practice number: Date of observation: Time of consultation: Duration of consultation: | |
| Technology | |
| What IT system(s) is being used | SystmOne: EMIS: Other: |
| Did the GP access the patient records before the consultation | Yes / No |
| What information was gained by accessing the patient record | |
| At what point does the GP access the computer | Prior to consultation: During consultation: Post consultation: |
| When are symptoms inputted onto the completer/patient record | Prior to consultation: During consultation: Post consultation: |
| Does the GP receive any interaction from the computer (alerts/pop-ups) | Yes / No |
| If alerts are received | What is the purpose of the alert?<br>What action was taken: |
| Referrals | |
| Were any referrals made during the consultation | No / Yes<br>To who: |
| When was the referral completed | During consultation: Post consultation: |
| What was the purpose of the referral | |
| How was the referral completed | Email: Telephone call: Online form: Through clinical system:<br>Template used? Yes/no |
| Professional collaboration | |
| Was there a discussion with another professional? | Yes / No |
| When did the discussion take place | During the consultation: Post consultation: Was the patient present: Yes / No |
| What was the purpose of the discussion | |
| Outcome of discussion | |

| | |
|---|---|
| Any other comments | |
| Patient engagement | |
| Did the patient report any symptoms | Yes / No |
| What symptoms were reported | |
| How are symptoms recorded | Coding: Yes / No Free text: Yes / No Other: |
| How were symptoms explored | Discussion: Yes / No Physical examination: Yes / No (If yes, give further detail below) |
| Were any physical observations completed | Blood pressure: Yes / No Heart rate: Yes / No SPo2: Yes / No Pulse: Yes / No<br>Listened to chest:<br>Other physical examination: Yes / No If yes, describe: |
| Was a new diagnosis given during the consultation | Yes / No If yes, what: |
| Were any symptoms of heart failure reported | Yes / No If yes what: |
| What heart failure symptoms were reported | Fluid retention: Yes / No Breathlessness: Yes / No Fatigue: Yes / No<br>Other: |
| If yes, what actions were taken | BNP test ordered: Yes / No ECHO ordered: Yes / No Specialist referral: Yes / No<br>If yes, to who:<br><br>Blood test: Yes / No Medication prescribed: Yes / No Name:<br>Dose:<br><br>Symptoms recorded on patient record: Yes / No Diagnosis given: Yes / No<br>No action taken: Follow-up review booked: Yes / No |
| If patient has existing diagnosis of heart failure | What previous diagnostic tests have been completed<br>BNP: Yes / No ECHO: Yes / No<br>Specialist referral: Yes / No To who:<br>Medication prescribed (which one): |
| Any other comments | |

**S2 Method: RAG rating for patients presenting with HF symptoms in primary care**

- Balance to be had between sensitivity / specificity of the main trigger – **NEED CONSENSUS AGREEMENT – TO CONSIDER the following prioritisation:**
    - 1 'Good' PLUS another (Moderate OR Poor) from another category OR 2 goods in the SAME category
    - 2 Moderate in EACH OF 2 Different Categories OR 3 moderates ACROSS 3 categories
    - 2 Poor in EACH OF 3 Different Categories OR 4 Poor ACROSS 4 categories

| | Poor | Moderate | Good |
|---|---|---|---|
| **1. Clinical history in Primary Care** | | | |
| Dyspnoea | | X | |
| ○ Exertional dyspnoea | | | X |
| ○ Orthopnoea | | | X |
| ○ Paroxysmal nocturnal dyspnoea | | | X |
| Fatigue/ Weakness | X | | |
| Swelling (Oedema): | | | |
| ○ Bilateral/dependent (esp. legs,<br>○ ankles, or feet/ sacrum) | | | X |
| Rapid or Irregular Heartbeat | X | | |
| Weight Gain | X | | |
| Risk Factor Assessment | | | |
| Cardiovascular Risk Factors: | | | |
| ○ Hypertension | | X | |
| ○ Diabetes | | X | |
| ○ Coronary artery disease | | X | |
| ○ Obesity | X | | |
| ○ Smoking | X | | |
| Previous Medical History: | | | |
| ○ History of myocardial infarction | | X | |
| ○ Valvular heart disease | | X | |
| ○ Arrhythmias | | X | |
| ○ Known cardiac conditions. | | X | |

| | | | |
|---|---|---|---|
| **Physical Examination** | | | |
| ○ Elevated blood pressure | X | | |
| ○ Arrhythmia | X | | |
| ○ Elevated jugular venous pressure (JVP) (? only right side/congestive?) | | X | |
| ○ Respiratory Crackles / Wheeze | X | | |
| ○ Displace apex beat | X | | |
| ○ S3 Gallop | | | X |
| ○ Other Murmurs | X | | |
| ○ Irregular rhythm | X | | |
| ○ Hepatomealy | X | | |
| ○ Ascites | X | | |
| ○ Dependent Oedema (Bilateral/ pitting/ sacral/ testicular) | | X | |
| **Initial Diagnostic Tests** | | | |
| Electrocardiogram (ECG): | | | |
| ○ Previous sign of myocardial infarction | | X | |
| ○ Left ventricular hypertrophy | | X | |
| ○ Arrhythmias | X | | |
| Blood tests | | | |
| ○ NT-proBNP elevated (Top – stand-alone marker) | | | X |
| Chest Xray | | | |
| ○ Cardiomegaly | | X | |
| ○ Pulmonary oedema | | | X |
| ○ Pleural effusion (bilateral only) | | | X |
| Echocardiogram | | | |
| ○ Top – stand-alone marker | | | X |
| **4. Medications** | | | |
| ○ On loop &/OR K-sparing diuretics | | | X |

**S3 Method: THINK-HF Coding Definitions**

**Risk Stratification System for Identifying Patients At Risk of Heart Failure in Primary Care**

The proposed system aims to identify patients at risk of heart failure in primary care by balancing sensitivity and specificity, using a combination of risk factors, symptoms, medications, and investigations. **The goal is to effectively flag high-risk patients without overwhelming clinicians with false positives.**

The main trigger will be driven by three filters (i) presence of two **risk factors** AND (ii) the presence of a **key symptom.**

**1. Main trigger**

**i. Primary Filter: Risk Factor-Driven Approach**

This system starts by evaluating **risk factors** first. A minimum of **two risk factors** is required to flag a patient for further assessment. The consensus on important risk factors includes:

| Risk Factor | Definition/Criteria |
|---|---|
| **Hypertension** | A QOF hypertension diagnosis code with no subsequent resolved code. This does NOT allow for latest raised readings (numeric) (without a code of Hypertension) |
| **Myocardial Infarction** | A code for MI at any time including proxy codes |
| **Diabetes Mellitus** | A QOF diabetes diagnosis code with no subsequent resolved code. Includes both Type 1 and 2 Diabetes |
| **Obesity** | Any BMI code that indicates the latest value >30Kg/m$^{2}$. Also includes finding/situation codes, eg 'Obese Class II / Body mass index 30+' |
| **Chronic Kidney Disease (CKD)** | The latest eGFR recording <60 mL/min with a previous recording <60 mL/min more than 90 days before the latest OR the latest CKD stage code recorded indicating CKD stage 3-5 |
| **Atrial Fibrillation** | An Atrial fibrillation diagnosis code with no subsequent resolved code |
| **Obstructive Sleep Apnoea** | A code for obstructive sleep apnoea. Includes mixed. Excludes central apnoea. Includes Paediatric codes also as linked to cardiomyopathy into adulthood |
| **Coronary Heart Disease (CHD)** | A QOF CHD diagnosis code. Includes CHD / Angina/ MI codes (allows for double weighting of MI) |
| **Valvular Heart Disease** | A code for valvular heart disease. Excludes MILD disease. Includes moderate/severe regurgitation/stenosis of ALL valves. Excludes Congenital |

| | |
|---|---|
| **Cardiomyopathies** | A code for cardiomyopathy including history of cardiomyopathy code. Widespread cardiomyopathy codes to include all potential including congenital / acquired cardiomyopathies |

**ii. Secondary Filter: Symptom Evaluation**

Once risk factors are identified, the presence of **key symptoms** will determine the next level of patient stratification. Patients must have at least **one key symptom** along with the risk factors to proceed with further investigation.

| Symptom | Definition/Criteria |
|---|---|
| **Orthopnoea** | At least one Clinical Finding code for ‘Orthopnoea (or its children)’ in the previous 5 years<br>*For Protocol trigger only: Includes free text entry (‘orthopnoea’). Excluding if ‘no’ before it |
| **Paroxysmal Nocturnal Dyspnoea** | Clinical Finding code for ‘Paroxysmal Noctural Dyspnoea (or its children)’ in the previous 5 years<br>*For Protocol trigger only: Includes free text entry (‘pnd’, ‘paroxysmal nocturnal dyspnoea’). Excluding if ‘no’ before it |
| **Peripheral Oedema** | At least one symptom code in the previous 5 years or a<br>Symptom Codes:<br>INCLUDES<br>- Feet/ankle/leg/thigh/lower limb<br>- Scrotum /<br>- Sacral<br><br>EXCLUDES<br>- Perineal (Penile/Vulval)<br>- Lymphoedema<br>- Lipoedema<br>- Myxoedema<br><br>For Protocol trigger only:<br>Includes free text entry (‘oedema’). Excluding if ‘no’ before it |

**iii. Exclusions:**

| Parameter | Definition/Criteria |
|---|---|
| **Age** | Under 16 years . |
| **Heart Failure** | Included in the QoF Heart Failure cluster without a more recent resolved code |
| **Heart Failure Excluded/Excepted** | Excluded/excepted in the past 12 months . |
| **Recent Trigger** | Excluded for 7 days after Pop-Up Alert launched |

**OVERALL (1→2→3→ Prompt for action) =** This algorithm highlights patients who are at risk, with at least one symptom, who have not had relatively recent ‘rule out/investigation’

**2. HF coding quality**

Heart Failure patients with coding quality issues should be flagged for review to ensure accurate coding.

| Coding quality issues | Definition |
|---|---|
| a. Heart Failure QOF Register but no QOF LVSD assessment | HF code at any time (without a subsequent resolved code) but no LVSD code recorded at any time. Includes QoF LVSD codes alongside other codes e.g., for a negative finding of LVSD, but shows it has been assessed. |
| b. Heart Failure QOF Register & LVSD Assessment but no Heart Failure Refined code | HF code at any time (without a subsequent resolved code) and a LVSD code but no code for Heart failure with reduced, midrange or normal ejection fraction |
| c. Heart Failure diagnosed but has a possible normal subsequent ECHO – Consider new HF code, HF Refined code or add resolved/exclude | HF code at any time with subsequent normal echocardiogram code (using codes Echocardiogram normal and Echocardiogram shows normal left ventricular function) |

**3. Codes suggestive of HF but no HF diagnosis**

Patients that are very likely HF but not coded should be flagged for review to ensure accurate diagnosis is coded.

| Patients that are very likely HF but not coded | **Definition** |
|---|---|
| a. LVSD codes but no Heart Failure diagnosis or exclusion codes | LVSD code (QoF) recorded anytime but no HF diagnosis code or no exclusion code within the last 12 months |
| b. Diastolic Dysfunction codes but no Heart Failure diagnosis or exclusion codes | Diastolic dysfunction code recorded anytime but no HF code any time or no exclusion code within the last 12 months |
| c. Heart Failure Proxy codes but no Heart Failure diagnosis or exclusion codes | HF proxy codes recorded anytime (history of/ management/ review/ plan etc) but no HF diagnostic code or exclusion code in the last 12 months |

**4. Diagnostic Testing**

For patients flagged based on **risk factors** and **symptoms**, specific diagnostic tests will provide further validation or guide clinical action.

| **Investigation based HF case finding** | **Definition** |
|---|---|
| a. Raised NT-proBNP but no referral to echo, echo result, or referral to cardiology from 2 weeks BEFORE up to 6 months AFTER the raised NT-proBNP . | - Latest NT-proBNP > 400 ng/L (including serum and plasma pro-brain natriuretic peptide level codes)<br>- No existing Heart Failure diagnosis<br>- No Heart Failure exclusion codes in the past 12 months AND AFTER the latest raised NT-pro-BNP<br>- No referral to echo, echo result coded, or referral to cardiology/HF team between 2 weeks prior to the latest raised NT-Pro-BNP result and 6 months after the result. |
| b. Raised NT-proBNP with the latest ECHO referral >6m ago and no ECHO result thereafter. | - Latest ever NT-proBNP is > 400 ng/L<br>- No existing HF diagnosis<br>- Latest Echo referral more than 6 months ago<br>- No echo result AFTER the latest ECHO referral |

| | |
|---|---|
| c. No NT-proBNP after symptoms code and has risk factors | - At least two risk factors (as defined above in i)<br>- A Heart Failure symptom (as defined above in ii) recorded in the last 5 years<br>- No subsequent NT-proBNP test. |

**5. Medications**

Patients on specific **heart failure medications** without a coded HF diagnosis should be flagged for review to ensure accurate coding.

| Medication | Definition |
|---|---|
| Entresto / Sacubitril and no Heart Failure diagnosis or exclusion codes | - Entresto / Sacubitril on current repeat template<br>- Issued in the past 3 months<br>- No current Heart Failure diagnosis<br>- No Heart Failure exclusion codes in the past 12 months |
| Hydralazine in combination with Nitrate but no Heart Failure diagnosis or exclusion codes | - Hydralazine in combination with Nitrate BOTH on current repeat template<br>- BOTH issued in the past 3 months<br>- No current Heart Failure diagnosis<br>- No Heart Failure exclusion codes in the past 12 months |
| Ivabradine but no Heart Failure diagnosis or exclusion codes | - Ivabradine on current repeat template<br>- Issued in the past 3 months<br>- No current Heart Failure diagnosis<br>- No Heart Failure exclusion codes in the past 12 months |
| SGLT2i but no Diabetes diagnosis or Heart Failure diagnosis or exclusion codes | - SGLT2i on current repeat template<br>- Issued in the past 3 months<br>- No current Heart Failure diagnosis OR current Diabetes diagnosis<br>- No Heart Failure exclusion codes in the past 12 months |
| On Digoxin and no AF diagnosis or Heart Failure diagnosis or exclusion codes | - Digoxin on current repeat template<br>- Issued in the past 3 months<br>- No current Heart Failure diagnosis OR current Atrial Fibrillation diagnosis<br>- No Heart Failure exclusion codes in the past 12 months |
| Loop Diuretics but no Heart Failure or exclusion codes | - Loop Diuretic on current repeat template (no time frame on issue)<br>- No current Heart Failure diagnosis<br>- No Heart Failure exclusion codes in the past 12 months |

| | |
|---|---|
| Loop Diuretics but no Heart Failure diagnosis or exclusion code AND abnormal NT-proBNP | - Loop Diuretic on current repeat template (no time frame on issue)<br>- No current Heart Failure diagnosis<br>- No Heart Failure exclusion codes in the past 12 months<br>- LATEST ever NT-proBNP is > 400 ng/L |
| Loop Diuretics but no Heart Failure diagnosis or exclusion code AND no history of NT-proBNP | - Loop Diuretic on current repeat template (no time frame on issue)<br>- No current Heart Failure diagnosis<br>- No Heart Failure exclusion codes in the past 12 months<br>- No NT-pro-BNP result EVER (whether raised or not) (numeric value) |

**S4 Methods: Baseline survey for HCPs**

UNIVERSITY OF LEICESTER

### GP Pre-study Feasibility Survey

#### Thank you for agreeing to complete our survey!

| |
|---|
| **TOPIC:** **Feasibility study to improve diagnosis of Heart Failure** |

**Please submit your survey by xx/xx/xxxx**

Pre-study Feasibility Survey_1_V1.0 30/10/23

#### GP Pre-study Feasibility Survey

* 1. Participant id number.

(This number is provided in your email. We will only use this number to link your surveys. All of your personal information will be kept pseudonymous).

* 2. What is your gender?

- ◯ Female
- ◯ Male
- ◯ Other

* 3. Which race/ethnicity best describes you? (Please choose only one.)

- ◯ White/White British
- ◯ Black/Black British
- ◯ Asian/Asian British
- ◯ Mixed race
- ◯ Rather not say
- ◯ Another race or ethnicity (please specify)

* 4. How long have you been qualified?

◯ Less than 5 years

◯ Between 5 and 10 years

◯ More than 10 years

* 5. As a GP do you have a special interest?

◯ Yes

◯ No

◯ Other (please specify)

6. If yes, what?

Interest 1

Interest 2

Interest 3

* 7. Have you ever used a clinical decision support system to facilitate assessment or management of patients?

☐ Yes

☐ No

☐ Don't know

8. If yes, what type?

☐ Electronic alert

☐ Paper based (e.g. check list)

☐ Other (please specify)

9. If yes, how helpful did you find it?

☐ Very helpful

☐ Somewhat helpful

☐ Neither helpful nor unhelpful

☐ Somewhat unhelpful

☐ Very unhelpful

10. What was good about it?

11. What was bad about it?

* 12. Have you ever taken part in training for the assessment and management of heart failure?

☐ Yes

☐ No

13. How recent was this?

☐ In the past year

☐ between 1 and 3 years ago

☐ between 3 and 5 years ago

☐ More than 5 years ago

* 14. Did your training sufficiently prepare you to assess and manage heart failure with reduced ejection fraction?

☐ Yes

☐ No

☐ Don't know

* 15. Did your training sufficiently prepare you to assess and manage heart failure with preserved ejection fraction?

☐ Yes

☐ No

☐ Don't know

* 16. Have you ever used a tool or resource to aid your assessment and management of heart failure during an encounter with a patient or as part of your training?

- ☐ Yes
- ☐ No
- ☐ Don't know

* 17. How confident are you in your knowledge of the NICE 2018 guidelines for <u>diagnosis</u> of heart failure?

- ☐ Very confident
- ☐ Fairly confident
- ☐ Not confident nor unconfident
- ☐ Fairly unconfident
- ☐ Very unconfident

* 18. How confident are you in your knowledge of the NICE 2018 guidelines for <u>treatment</u> of heart failure with <u>reduced ejection fraction</u>?

- ☐ Very confident
- ☐ Fairly confident
- ☐ Not confident nor unconfident
- ☐ Fairly unconfident
- ☐ Very unconfident

* 19. How confident are you in your knowledge of the NICE 2018 guidelines for <u>treatment</u> of heart failure with <u>preserved ejection fraction</u>?

- ☐ Very confident
- ☐ Fairly confident
- ☐ Not confident nor unconfident
- ☐ Fairly unconfident
- ☐ Very unconfident

* 20. Of the patients you have seen with heart failure or you thought were at risk of heart failure, with what proportion did you implement the NICE guidelines?

- ☐ All
- ☐ Most
- ☐ Some
- ☐ A few
- ☐ None

21. Where you didn't implement the NICE guidelines, can you list some of the key reasons?

| |
|---|
| Reason 1 |
| Reason 2 |
| Reason 3 |
| Reason 4 |
| Reason 5 |

* 22. Please indicate below whether you are willing to continue to participate in this study

◯ Yes, I am happy to continue to participate in the study

◯ No, please don't send me any further surveys

**GP Pre-study Feasibility Survey**

**Thank you for completing  our survey!**

There is one further survey at the end of this feasibility study.

The next one will be a lot shorter.

**S5 Methods: Post study survey for HCPs**

UNIVERSITY OF LEICESTER

### GP Post-study Feasibility Survey

#### Thank you for agreeing to complete our survey!

| |
|---|
| **TOPIC: Feasibility study to improve diagnosis of Heart Failure** |

**Please submit your survey by xx/xx/xxxx**

Pre-study Feasibility Survey_1_V1.0 30/10/23

* 1. Participant id number.

(This number is provided in your email. We will only use this number to link your surveys. All of your personal information will be kept pseudonymous).

[ ]

* 2. How helpful did you find the training on using the THINK-HF clinical decision support system?

- ☐ Very helpful
- ☐ Somewhat helpful
- ☐ Neither helpful nor unhelpful
- ☐ Somewhat unhelpful
- ☐ Very unhelpful

3. What was good about it?

[ ]

4. What was bad about it?

[ ]

* 6. How helpful was the THINK-HF clinical decision support system in prompting you to tests for heart failure?

- ☐ Very helpful
- ☐ Somewhat helpful
- ☐ Neither helpful nor unhelpful
- ☐ Somewhat unhelpful
- ☐ Very unhelpful

* 7. How helpful was the THINK-HF clinical decision support system in helping you diagnose or rule out heart failure?

- ☐ Very helpful
- ☐ Somewhat helpful
- ☐ Neither helpful nor unhelpful
- ☐ Somewhat unhelpful
- ☐ Very unhelpful

8. Overall, what was good about 'THINK-HF'?

9. Overall, what was bad about 'THINK-HF'?

10. How confident are you in your knowledge of the NICE 2018 guidelines for diagnosis of heart failure?

☐ Very confident

☐ Fairly confident

☐ Neither confident or unconfident

☐ Fairly unconfident

☐ Very unconfident

* 11. How confident are you in your knowledge of the NICE 2018 guidelines for treatment of heart failure with reduced ejection fraction?

☐ Very confident

☐ Fairly confident

☐ Neither confident or unconfident

☐ Fairly unconfident

☐ Very unconfident

* 12. How confident are you in your knowledge of the NICE 2018 guidelines for treatment of heart failure with preserved ejection fraction?

☐ Very confident

☐ Fairly confident

☐ Neither confident or unconfident

☐ Fairly unconfident

☐ Very unconfident

13. Of the patients that you were alerted to by 'THINK-HF', with what proportion did you follow the prompts?

- ☐ All
- ☐ Most
- ☐ Some
- ☐ A few
- ☐ None

* 14. If not, why not?

15. If you didn't respond to the prompts, can you list some of the key reasons why?

| |
|---|
| Reason 1 |
| Reason 2 |
| Reason 3 |
| Reason 4 |
| Reason 5 |

**GP Post-study Feasibility Survey**

**Thank you for completing our survey!**

**S6 Methods: Interview guide for HCPs.**

**RECORD OF INTERVIEW FOR STUDY PARTICIPANTS.**

**Name of Interviewer:** ________________________________________

**Name of others present:** ________________________________________

**PARTICIPANT ID No:** ________________________________________

**PRACTICE ID No:** ________________________________________

**SEX:** ________________________________________

**YEAR OF QUALIFYING:** ________________________________________

**CURRENT JOB ROLE:** ________________________________________

**YEAR OF QUALIFYING:** ________________________________________

**TIME IN GENERAL PRACTICE (YEARS):** ________________________

**EXPERIENCE WORKING IN A CARDIOLOGY SETTING** YES/NO

YEARS__________________

#### INTERVIEW GUIDE FOR HEALTHCARE PROFESSIONALS

**Overall aim of the process evaluation interviews:** To understand what worked, what didn't work, for whom and why in diagnosing heart failure, and improving health outcomes for heart failure patients, and to inform future implementation.

**Introduction to the healthcare professional**:

*Hi, I am [name] and I'm from [organisation]. Thank you for taking part in this interview. As discussed, we are trying to find out how we can improve diagnosis of heart failure. You can change your mind about talking to me at any time before or during the interview and stop the interview at any time. Are you happy to continue? [If no, thank them for their time and end interview; if yes continue.] Thank you [name] for agreeing to take part.*

*We are interviewing health professionals involved in the care of heart failure patients. We will use your feedback and the feedback of others to write a summary of what people have told us. We ask that you do not name any patients in our interview when discussing the care you provide. Are you happy for me to record the interview? I will keep the file in a secure location until we finish writing the report and then it will be destroyed. Our interview will be transcribed professionally and we will ensure your privacy and confidentiality.*

*Do you have any questions before we start?*

*Note: Key questions in bold, with probing questions in non-bold. Questions do not have to be asked in this order, and not all questions have to be covered.

| **Domain 1: To understand the context of the site and health system, and how diagnosis of heart failure patients fits within this context.** |
|---|
| **Warm up:** What is your role? Can you describe your role in the 'THINK-HF' feasibility study? How did you get involved and why?<br><br>**What do you think are the main purposes of the THINK-HF feasibility study?** [Probe: increase awareness of risk indicators, tailored information, prompts to appropriate tests].<br>• What types of patients are challenging to diagnose with heart failure? Why is that?<br>• Are there any health service factors that make heart failure diagnosis difficult?<br>• How important is it to diagnoses heart failure promptly?<br><br>**What is the assessment and diagnosis pathway for patients presenting with heart failure symptoms or signs at your practice?** [probe: BNP test, echo, referral to specialist]<br><br>**What is the treatment pathway for patients with heart failure at your practice?** [probe: prescribed drugs, specialist review, care plan, follow-up] |

**What do you think are the main causes of death or hospital admissions within 1 year for heart failure patients? Are they preventable in any way?** [probe: patient level factors such as MLTCs, lack of knowledge, adherence or access to care; health care level factors such as access to specialist services]

**<u>Domain 2: To understand the implementation barriers and facilitators</u>**

**Have you used clinical decision support systems previously? If yes, did you find them helpful? Why, why not?**

**What in your perspective has gone well in using 'THINK-HF'? What in your perspective has been difficult? Why is that? Can you provide some examples?**

- Did you find the prompts helpful? Why, why not?
- Did you respond to the prompts? If not, why not? Can you provide some examples?
- Did you adapt the implementation in anyway?

**What did you think about the training session on 'THINK-HF', was it adequate?**

**What do you think of the adoption of its use from the patients' and health providers' perspective?**

- Is it adequate or not, and why? Can it be improved?
- Was there an opportunity to improve its implementation?

**Were there any health system issues that impacted upon the implementation? [**probe about: workforce, health financing, governance, fitting in with current information technology such as EHR]

**<u>Domain 3: To understand the outcomes and future implementation</u>**

**Do you think this trial will be successful? Why is that?**

- Would it improve time to heart failure diagnosis?
- Will it reduce the number of patients diagnosed in hospital?
- Will it improve the recognition and coding of heart failure?
- Upon hindsight, is there anything that you would do differently?

| **Would it be beneficial and feasible to continue and embed 'THINK-HF' into usual care?**<br>• Do you think this will be widely accepted by GPs in their practice? Why or why not?<br>• How do you think general practitioners could be best supported in diagnosing heart failure? Do you see 'THINK-HF' playing a part in this process? |
| --- |
| **Concluding question and Statement** |
| Is there anything else you would like to say that we have not talked about in this interview?<br>Thank you so much for your time and for sharing your insights. |

**RESEARCHER FIELD NOTES:**

How do you think the interview went?

What struck you as important?

What further questions/areas would you like to explore in the next interview?

**S7 Method: Interview guide for patients.**

**<u>RECORD OF INTERVIEW FOR STUDY PARTICIPANTS.</u>**

**Name of Interviewer:** ____________________________________________

**Name of others present:** ____________________________________________

**PARTICIPANT ID No:** _____________________________________________

**PRACTICE ID No:** _____________________________________________

**AGE:** _____________________________________________

**SEX:** _____________________________________________

**MEDICAL HISTORY** _____________________________________________

**REASON/S FOR CONSULTATION** _______________________________________

**INTERVIEW GUIDE FOR PARTICIPANTS**

**Overall aim of the process evaluation interviews:** To understand what worked, what didn't work, for whom, and how to improve consultation experience for patients presenting with common symptoms (breathlessness, ankle swelling and tiredness), and to inform future implementation.

**Say to the participant**:

*Hi, I am [name] and I'm from [organisation]. Thank you for taking part in this interview. As discussed, we would like to ask you questions about your experience of your GP appointment and also in relation to assessment and diagnosis of your medical condition/s. You can change your mind about talking to me at any time before or during the interview and stop the interview at any time. Are you happy to continue? [If no, thank them for their time and end interview; if yes continue.]*

*We will use your feedback and the feedback of others to write a summary of what people have told us. There will be absolutely no identification of any real names or identification of where you live or which health professionals you have seen. Are you happy for me to record the interview? I will keep the file in a secure location until we finish writing the report and then it will be destroyed. Our interview will be transcribed professionally and we will ensure your privacy and confidentiality.*

*Do you have any questions before we start?*

*Note: Key questions in bold, with probing questions in non-bold. Questions do not have to be asked in this order, and not all questions have to be covered.

| **Domain 1: To understand the patient context and usual care** |
|---|
| **Can you tell us about your recent GP appointment with breathlessness/ ankle swelling/ tiredness (delete as appropriate)?**<br>• When did your symptoms start?<br>• How do the symptoms described affect your daily life?<br>• How have they been managed?<br><br>**During your appointment, what were some key things that you were advised to do? How was it to follow this advice? What went well or not?**<br>• Have you been told what may be causing your symptoms?<br>• What advice were your given? |
| **Domain 2: Intervention implementation and mechanisms**<br>***As you know, we are exploring whether an electronic intervention that the GP is using to help diagnosis is helpful. The next questions will explore this care you received in greater detail. Please feel free to be honest about what it was like for you, as any feedback you provide will help us improve the care we provide.*** |

**Did the GP mention that he/she was using a symptom diagnosis tool?**

- Did the GP ask you about conditions (such as lung problems, high blood pressure, rhythm problems, heart problems, angina [chest pain])
- Have you been referred for any tests or been referred to a specialist in relation to your symptoms?

**What did you like about your appointment?**

- Did your appointment help you to understand what might be causing your symptoms and how you could manage them? Can you provide an example of how that happened?
- How satisfied were you with the information given to you during your appointment?

**What didn't you like?**

- Could you provide an example of what happened?

**What else would you liked to have changed about your appointment?**

- Was the information given to you helpful? Why/ Why not?

**<u>Domain 3: Exploring more about mechanisms of the patients' outcomes</u>**

**Do you think diagnosing the cause of symptoms earlier will help improve your health?**

- Did you have any follow up after your appointment? What did you like/dislike about follow up?
- Can your health providers do more for you? In what way?

**How are you managing now?**

- Have you been to see your GP again in the past two weeks? Why was that?
- How are you managing to keep well, and manage your daily activities? (probe: self-care, and other supports)
- What has been difficult? What has been going well?

**Concluding question and Statement**

Is there anything else you would like to say about your recent GP appointment that we have not talked about in this interview?

| |
|---|
| Thank you so much for your time and for sharing your insights. |

**RESEARCHER FIELD NOTES:**

How do you think the interview went?

What struck you as important?

What further questions/areas would you like to explore in the next interview?

### Supplementary Tables

**S1 Table: Process evaluation quotes: GPs**

| Domain | Representative quotes |
| --- | --- |
| **Reach** | • *"It was particularly relevant for me … I had a patient with liver disease, and your pop-up very helpfully came on at that point and reminded me not to forget the heart." ……"Multi-comorb patients … like a patient with decompensated liver disease … your pathway reminded me that I should not forget the heart." (ID 21)*<br>• *"If someone presents to me with swollen feet then generally I will check a BNP … it fits with what we should be doing, but it highlights the areas where we maybe aren't." (ID 21)*<br>• *"It's [THINK-HF aim is] identifying new cases [of HF] … there are a significant number of people who are probably under-diagnosed." ……… "Very obese patients can sometimes be very difficult to differentiate … whether in fact they have got heart failure." ………. "Language can be an issue … if people can't describe their symptoms very accurately … you may not think of heart failure." (ID 29)*<br>• *"Those patients who have multi-comorbidities… COPD in particular… the assumption might be that it's the COPD… the heart failure might not come to a clinician's mind straight away." (ID 30)*<br>• *I think it's useful to find patients with breathlessness or swollen legs who have never been considered have they got heart failure or not, and then we can get the ball rolling with investigating, so BNP, echo, and then confirm or rule out." (ID 21)* |
| **Effectiveness** | **Alerts**<br>• *"It reminded me that I should not forget the heart… especially in multi-morb presentations." ……… "Liver problems can also cause shortness of breath and oedema… but your pop-up very helpfully came on… It sort of reminded me not to forget the heart… I can't thank you enough for that liver patient that you prompted." (ID 21)*<br>• *"The one that it did, it was actually relevant, cause the guy had come in with a swollen leg… This pop-up did help for me to look back and recognise that… he's presented with this swelling over these last couple of months… it may still be helpful to do the bloods and check the BNP as well as assessing for the DVT." (ID 20)*<br>• *"It's almost every single day, …. that I have patients with worsening shortness of breath or your pedal oedema, chest pains and things, which can all be cardinal signs of heart failure…..and it has alerted me to think about heart failure. I* |

*did feel your study was very useful as a sort of a visual reminder to not forget considering these, considering heart failure as a differential in these patients." (ID 21)*

**Prompts**

- *"The prompts I found one of the most helpful things… it's simple, it's effective, it gets the message across." (ID 19)*
- *"If it's a relevant patient you would respond to the prompt… you are going through the whole pathway and making sure you're not missing out on things." (ID 31)*
- *"If you know the right type of heart failure you can… pitch the right type of support… medications… specialist support… both from a prognostic and a management point of view." (ID 21)*
- "I had not thought of heart failure and it was very good to have that prompt." (ID 21)
- *"If I've gone in for a consultation and it's come up, I think probably I would say 70 to 80% of the time I've followed through it, because it is relevant and it's related to my consultation." (ID 21)*
- *"we have got people in for blood tests, or said oh, you had a BNP but never had an echo, then we've looked into that." (ID 22)*
- *"it's quite clear from the template, if you go into it, at what stage they are in the process and what the next step might be, and then we can arrange that." (ID 22)*
- *"The prompts I found one of the most helpful things. It delivers, it makes you think of heart failure. That's all it says, and I think that's just what the intention is, just to trigger that neuron, have you thought of it? If not, it might be there's something there. It's simple, it's effective, it gets the message across. Yeah, that was great." (ID 19)*
- *"if you've got a busy clinic and you're running behind, as I often do as well, then it just helps to remind me." (ID 20)*
- *"if it's a relevant patient you would respond to the prompt, so yeah, definitely, so you're gonna go through it." (ID 31)*

**Diagnosis & outcomes**

- *"We'll get more people diagnosed and on the correct treatment, thereby reducing emergency admissions." (ID 22)*
- *"Coding has always been one of the difficulties with heart failure… that is on our list to do… because coding has always been one of the difficulties." (ID 30)*
- *"With this one, we can code it… once we've done the echo… it triggers us to… do the specific coding." (ID 31)*
- *"Should reduce hospital diagnoses by picking up earlier." (ID 29)*
- *"You can pre-empt some admissions by adequately managing the patient and helping them understand what to expect." (ID 29)*
- *"But I think from a patient point of view it's safe and it's for enhanced patient care, so I think it's a good thing." (ID 21)*

- *"We're doing a disservice by a late diagnosis, just like similar to cancer… missed opportunities… to make sure they're getting access to the treatments… as early as possible." (ID 20)*
- *"I think it's very important, as later diagnoses do lead to increased adverse outcomes". (ID 19)*
- *"Early diagnosis can help prevent progression and manage avoidable risk factors." (ID 29)*
- *"if you diagnose early you can educate patients to see what lifestyle changes they can make, we can make appropriate referrals to secondary care or heart failure nurses, make sure they're on optimal medication to slow progression, reduce emergency admissions" (ID 21)*
- *"If it's delayed then they're gonna be going in and out of hospital, so it's gonna have a big impact on the patient and NHS costs and everything." (ID 21)*
- *"I think if they're diagnosed early and started on the correct treatment and monitored properly then that's got to reduce the risk of emergency admissions and death" (ID 22)*

**Behavioural change**

- *"It [THINK-HF] changes the way we think about diagnosing disease at an early stage."* (ID 19)
- *"Makes you think HF ... simple, effective." (ID 19)*
- *"certainly led to change in practice and more recognition and more alertness towards heart failure" (ID 21)*
- *"there was a drive two or three years ago….. to increase coding [of heart failure], and making sure that if you had heart failure you were coded as having heart failure…..I was aware that there were gaps in our coding and there were perhaps people with heart failure that weren't coded and therefore weren't on the correct treatment" (ID 21)*
- *"As a clinician in the consultation with the patient I may not always be thinking so broadly in terms of changing outcomes as patients present with symptoms we're primarily focused on the symptom at the time, and this I think increases or changes the way we think about diagnosing disease at an early stage."* (ID 20)
- *"They were [management] into this idea and they've all been very encouraging of us to do it. It's something that we've already been doing, but it's now a more formalised, structured method of doing."* (ID 21)
- "*I think it has raised, in a general sense, awareness and cognisance of heart failure in the organisation…..we're starting to, I hope, think a little bit more proactively*." (ID 19)

**Adoption**

**Decision support; what has gone well?**

- *"It clearly works, it does pop up … it integrates well."* (ID 22)
- *"Younger doctors do tend to use them [decision-support tools] more."* (ID 29)
- *"Sometimes it can be a bit excessive … but 90–95% of the time it's very useful." (ID 21)*
- *"If I've gone in for a consultation and it's come up, probably 70-80% of the time I've followed through it." (ID 21)*
- "*I think it's completely appropriate. It's something that's needed, it has identified a healthcare need"* (ID 19)
- "*I've not had any feedback from anyone about that popup being annoying, frustrating or any issues with it. And you know we do have it for others, so I guess no feedback means it's OK!" (ID 20)*
- *"the fact that you can request ICE and stuff, the results through, the bloods can be requested through there, that was all helpful as well, so I think that integrates really well" (ID 20)*
- *"I think that it's a really good effort, and I hope actually it does increase the awareness amongst clinicians, because we're the ones, like all different types of healthcare professionals, coming into contact with these patients and to be thinking of this" (ID 20)*
- *"I guess ultimately it's, at the moment it pops up in the relevant cases, so yeah, it seems to be integrated and working well." (ID 30)*
- *"So I think this [THINK HF] is actually really good, because it helps you go through the actual diagnosis systematically, so I think it links in quite well actually." [ID 31)*
- *"It is quite easy to use. I mean like I say, it's easy to use, you're going through it, you click what you need to. So yeah, and you can put the codes in as well, so I think generally it's good." (ID 31)*
- *"I think it's really useful before you even think about referring. Yes, we can refer, so we always have that, but I feel like it makes you a little bit more independent in your diagnoses, and then you have a little bit more information before you do that referral. " (ID 31)*
-

**Decision support; what has not gone well?**

- *"It certainly is very sensitive … That's a good thing, but then you do get the alert fatigue issue." (ID 22)*
- *"There's something about software… the amount of pop-ups… which does cause a little bit of fatigue of the mind." (ID 19)*
- *"I'm probably one of the ones who closes them down and just does my own thing … too old in the tooth!" (ID 29)*
- *"Because the time is limited and how things are so busy, what I tend to use is I use it just as a reminder, and not actually use, go into the tool too much myself, but look at the patient, screen them, and then move forward." (ID 36)*

- *"Integrated OK I think, but with a lot of these prompts and pop-ups, a lot of people feel that it's annoying sometimes, and then the automatic reaction is to switch off and do what you were already doing. It could be anything, it could be any kind of pop-ups, because there's so much of things bombarding you when you are in that 10-minute consultation." (ID 36)*

**Implementation** **Training:**

- *"she spoke to all of us and spent time with us, and also told us what to expect, the pop-up tool and things like that. That made it very informative and yeah, it was quite good". (ID 21)*
- *"The training session was very good. It was clear to me how to use the tool and what to expect." (ID 19)*
- *"It was very helpful to have the online video, which was just a few minutes of demonstration. That's very helpful, but in a large healthcare practice there's another layer of trying to get everyone to use that, have a look and engage. ..,. The content was great, but it's how to roll out the content to clinicians who are already busy receiving many updates from different parts of the NHS." (ID 19)*
- *"I think having that initial meeting and discussion, that tutorial almost on it, was really useful, because sometimes just having to read through the literature sometimes is less effective, so actually having a demo of the tool was quite useful, actually seeing it in action before using it" (ID 30)*

**Facilitators:**

- *"implementation-wise, again it was great, just a sort of video demonstration of this is how it comes up, that would be useful, but otherwise I don't think you can do anything different." (ID 21)*
- "*Think perhaps just a closer check-in and some guidance or a plan to be created with a senior clinician like myself…….. In hindsight I think I could have planned more, done more, to induce the rest of the team, make them aware." (ID 19)*
- *"Just the usual things, isn't it, just making sure everybody knows about it, and reminding people about it and explaining the background behind it and why it's there. So to just maintain that clinical awareness and things for it, so that's the biggest thing." (ID 20)*
- *"But having it [THINK-HF] added into especially locality meetings and things that take place, as a reminder for everybody, I think that's the most important bit." (ID 20)*
- *"So some of them are very much intrinsically linked with national guidance for example, so those ones are incredibly useful." (ID 30)*

- *"Because we've used similar, not in relation to heart failure, but similar tools in the past, where those pop-ups happen pretty much for every single patient, and then you end up ignoring it…….I think with this tool actually it does seem to have been quite specific. So it's only actually popping up, from what I've seen so far, for actually quite relevant cases. So it's not overwhelming us, it seems to be well set up in identifying more specific cases. (ID 30)*
- *"If it can be like a gentle nudge, I think people will be more accepting it, whereas if it's too much on your face then people don't want to spend that, let's say extra few minutes, whatever. Because I mean as it is, the working life, it's so busy. So in the right context, in the right patients, I think as a gentle reminder, that that would be useful I think, yeah." (ID 36)*

**Barriers:**

- *"When you've got 10 minutes per patient and they come in with a list of things … that's the only limiting factor." (ID 21)*
- *"A patient typically will present with a symptom… allocated into an appointment slot which may not be enough time in the first place." (ID 19)*
- *"There's something about software, the amount of pop-ups and different providers, which does cause a little bit of fatigue of the mind, and it may feel unhelpful at times if our clinical thoughts are in one direction and then there's a tool saying something different. But ultimately it is helpful, yes." (ID 19)*
- *"I think sometimes it can be a bit excessive, …. But it's generally very useful most of the time, I'd say probably 90, 95% of the time it's very useful." (ID 21)*
- *"Those ones where I do cancel the template … it leaves a footprint on the patient's record, which is a little bit annoying." (ID 22)*
- "*The only challenge was, for whatever reason you go into the patient's records, the pop-up would come up, and when you mark it as sorry, I've not gone in for heart failure sort of thing, I've gone in to manage their infection or whatever, says protocol not followed …..So I think every entry sort of leaves a sort of footprint, doesn't it, or a record. That was probably the only thing that was a little bit of a downer for me, but otherwise I don't think that there was any other problems that I come across. (ID 21)*
- *"I only thing I can think of is time, when they've got 10 minutes per patient and they come in with a list of things, yeah, and if heart is not their top priority, but it's your agenda, so how [inaudible 00:09:35], that's the only limiting factor I suppose" (ID 21)*
- *"The system is not set up for asymptomatic patients or patients with low-grade symptoms to proactively be reviewed. Things are changing, but it's not set up that way." (ID 19)*
- *"My personal feeling is the same change may not have happened with other clinicians… who are more distant to this particular piece of research and development." (ID 19)*

**Adapted implementation:**

*Task sharing*

- *"On probably one or two occasions I may have sort of deferred it to… I think you get a task, don't you, to remind yourself to go back in and sort it out later, so I've done that." (ID 21)*
- *"Rather than deal with the pop-up in clinic, we collect them and refer to our research administrator, who then puts them in a clinic for me to look at … it's probably good to do them all in one go…….I've done a couple of clinics like that probably about fifteen patients so far … it's been very useful." (ID 29)*
- *"The routine reviews would be done by the practice nurses, so they can assess for any change in symptoms and then they can let the doctors know if there are any concerns." (ID 21)*
- *"if the patient is coming in to see our nurse as part of their long-term condition review ….I do think that there is value, if it comes for the nurse as well, because if the patient then starts talking to them about something else, then hopefully that's triggered and prompted them. Oh, they said breathlessness, and there was that prompt that came up, and maybe I need to send this task on to a doctor." (ID 20)*
- *"we also find that it's probably good to do them all in one go, because you're in that heart failure mode as it were, and then you can look through their records, see if the pop-up is appropriate, whether the diagnosis has already been made and they're already on management, or whether they need some further investigations." (ID 29)*
- *"So what we've done is we then send them over to another inbox, and one of my colleagues is then actually looking through them and going into those patients in almost a separate environment away from that initial clinical contact, if that makes sense." (ID 30)*
- *"There's certainly benefit… the extent of it I'm not sure. Different parts are useful for different staff… some will use the templates, others the alerts only." (ID 19)*

*Continued support*

- *"There's perhaps something there at a network level… dedicated funding for four or five hours of clinician time for someone to log in to all the practices… and start helping through using the searches." (ID 29)*
- *"In-house at the practice we needed someone I think to increase awareness and training with the staff, to ensure it's used to its fullest extent." (ID 19)*

*Proactive Searches*

- *"And I'm aware that in the background members of my team are running searches, and I find that side of it more helpful, … they'll give me a list saying right, these people have got coded, never had a BNP….. And I'll go through the list." (ID22)*
- *"I have found it more helpful running searches rather than responding to the prompts." (ID 22)*
- *"The redirecting is just seconds, cause all you're doing is pinging a message to the administrator to tell them this patient has had a pop-up and can someone look at it basically." (ID 29)*

**Maintenance** **Were health system factors affecting implementation of THINK-HF.?**

- *"So whether you want to, for example, do a BNP for a patient, obviously in primary care it's not gonna be done on the same day, so blood tests to come back, and also echocardiograms, they're gonna take some time for the patient to have it and for the report to come back. So I think those are the major obstacles"* (ID 32)
- *"I think if the echo becomes more readily available, that would be a good thing. If you can get it faster." (ID 32)*
- *"Well one of the things we find is the quality of the results we get, for example echocardiogram, I don't know whether you have… I mean you can't do anything here, but there are various providers doing the echocardiogram, and the quality, the standards are probably not up to the mark I think, you know?" (ID 36)*
- *"Yeah, it'd be good if we could get echoes quicker." (ID 22)*
- *"I guess it would just be probably patient access. I mean it's not directly limiting our ability to diagnose, but patients nationally find it difficult to access the GP, …. they're often maybe put off to contact the GP over quite mild symptoms, because they think, or they perceive, that getting an appointment will be really difficult….., (ID 30).*
- *"I think it's got better for the fact that we have the community access to our echocardiogram requests. Think usually once we put the request in, I think patients get their scans within about four to six weeks, and there's multiple sites in the community. So I think that's made a huge difference…… The blood tests, so this'll be practice dependent and obviously things like staffing, sickness…..So getting that initial BNP, which often you'd want that first before requesting the echo, it feels like it could take longer than I'd usually like, but it's usually done within a month. And in the grand scheme of it, given the patients aren't acute, then I think it's acceptable." (ID 20)*

**Do you think the THINK-HF trial will be successful?**

- *"I think it works, and I think the more we're aware of it and less scared of it, or annoyed by it, I think we will act on it more. So I think it should be something that the more we get used to it the more we actually use it. The use should grow I think.." (ID 22)*
- *"It's overall very positive … it made us all think about heart failure a bit more." (ID 21)*

- *"Mmm, yeah, yeah, give yourselves a pat on your back!  Yeah, I can't thank you enough for that liver patient that you prompted.  I think it was very useful to learn, it's a great thing." (ID 21)*
- *"I think it'll be a useful tool … how practices manage it, whether they do it like us or differently, will be interesting." (ID 29)*
- *"There's certainly benefit… the extent of it I'm not sure. Different parts are useful for different staff… some will use the templates, others the alerts only." (ID 19)*
- *"There's scope for good work and improvement if the Think-HF team finds a way to collaborate with [Hospital] and primary care networks." (ID 19)*
- *"You guys should probably share your numbers with us as to how many new diagnoses you've had." (ID 21)*
- *"Keeping CPD up to date is probably the most appropriate way to support GPs [in the diagnosis of heart failure]." (ID 29)*
- *"I think so.  I think it would be a really good tool actually, because we don't have one that's as thorough as this*. " (ID 31)

**S2 Table: Process evaluation quotes: Patients**

| Theme | Relevant Quotes |
|---|---|
| **Access, routes & gatekeeping** | • *"You're 29th in the queue… there's no appointments." ID43*<br>• *"you have to do is ring at eight o'clock every morning to make an appointment. And if you can't get an appointment then I've ruined five hours of sleep." ID12*<br>• *"I've called at eight o'clock before, and I can get through at like 10 past, quarter past eight, and all the appointments are gone. And you can't prebook either." ID53*<br>• *"the online form is from eight o'clock onwards, and if you submit it later then appointments are not available……otherwise you're seen by emergency clinics, and they don't know you, they don't have your history, and then you've got to go through everything again and that's really frustrating," ID51*<br>• *"I quite like telephone conversations, cause they're much easier for me to deal with." ID24*<br>• *"I know they want you to use the online service… from a sight point of view it's not always an easy system for me to navigate." ID27* |
| **Continuity & relationship work** | • *"I'd like to see the same GP… it's a little bit frustrating when you have to answer the same question nine times." ID24*<br>• *"it was a nurse practitioner that I saw. I mean I think the last three times I've been I've seen a nurse, not an actual GP. Can't remember the last time I saw a GP!" ID10*<br>• *"my normal GP, I trust him implicitly, and he's the one that if I ask him a question he tells me how it is. So I respect him greatly on that, and if I can get to see him I prefer to see him." ID11*<br>• *"I very rarely see the same doctor twice, it's been increasingly more difficult since Covid-19, thereabouts." ID14*<br>• *"I try and see the same doctor……. that's not always practical as well, because they're not available or they don't do clinics on certain days. And again you're having to explain everything to different doctors then…….. it's just too much to explain everything again and again and again." ID51*<br>• *"I have community mental health worker, nurse to come and see me, and she makes 30-minute appointments with the GP, because you never used to get the same GP……. and she's sorted it out now." ID50* |

| | |
|---|---|
| **Communication, explanation & follow-up** | • *"I was supposed to have a follow-up appointment… and I haven't heard anything. I'm still waiting." ID48*<br>• *"They give you that much information… sometimes it would be good for them to write things down." ID53*<br>• *"It would have been nice to think they could investigate further." ID10*<br>• *"I kind of think I got all the answers that they could give me, if that makes sense. I feel like the system is not geared up as well as it could be for heart patients……… I just get the feeling that they're in the moment and there's not a path to go down, that there's no plan." ID14*<br>• *"sometimes they give you that much information, and they don't write things down, they tell you these fancy names and you don't understand it." ID53*<br>• *"in my discharge letter it said NSTEMI. No-one explained to me what an NSTEMI was. I googled it to find out what it was". ID51*<br>• *"[HF nurse] arranged for me to be seen… it proved a marvel in terms of the swelling." ID54* |
| **Delays and misattributed diagnoses** | • *"It was at least 14 years before I was actually diagnosed [HF]." ID38*<br>• *"I thought it was just like an anxiety attack… but it turned out it was cardiomyopathy." ID39*<br>• *"To be listened to… having the tests done earlier would have helped" ID39.* |
| **Practical access barriers & digital processes** | • *"Because of my mobility issues… it's made it very difficult for me to actually get there." ID24*<br>• *" I know they want you to use the online service …. from a sight point of view it's not always an easy system ……... I think a part of it is my age and my being a technophobe, and part of it is the visual thing, that they're not always as receptive to my need for reasonable adjustments as I think they could be." ID27* |

### Supplementary Figures

#### S1 Figure: Heart failure flag on clinical record

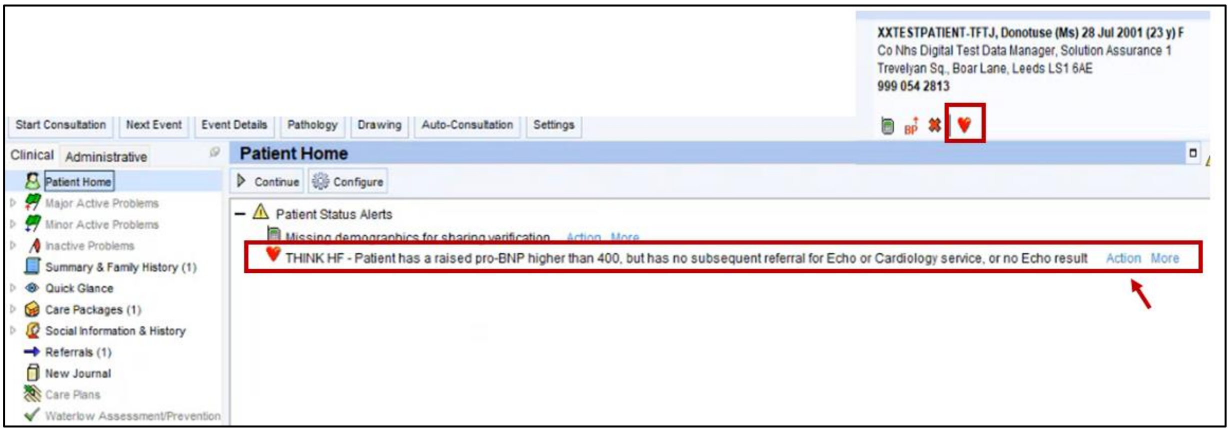

#### S2 Figure: Clinical template for coding issues

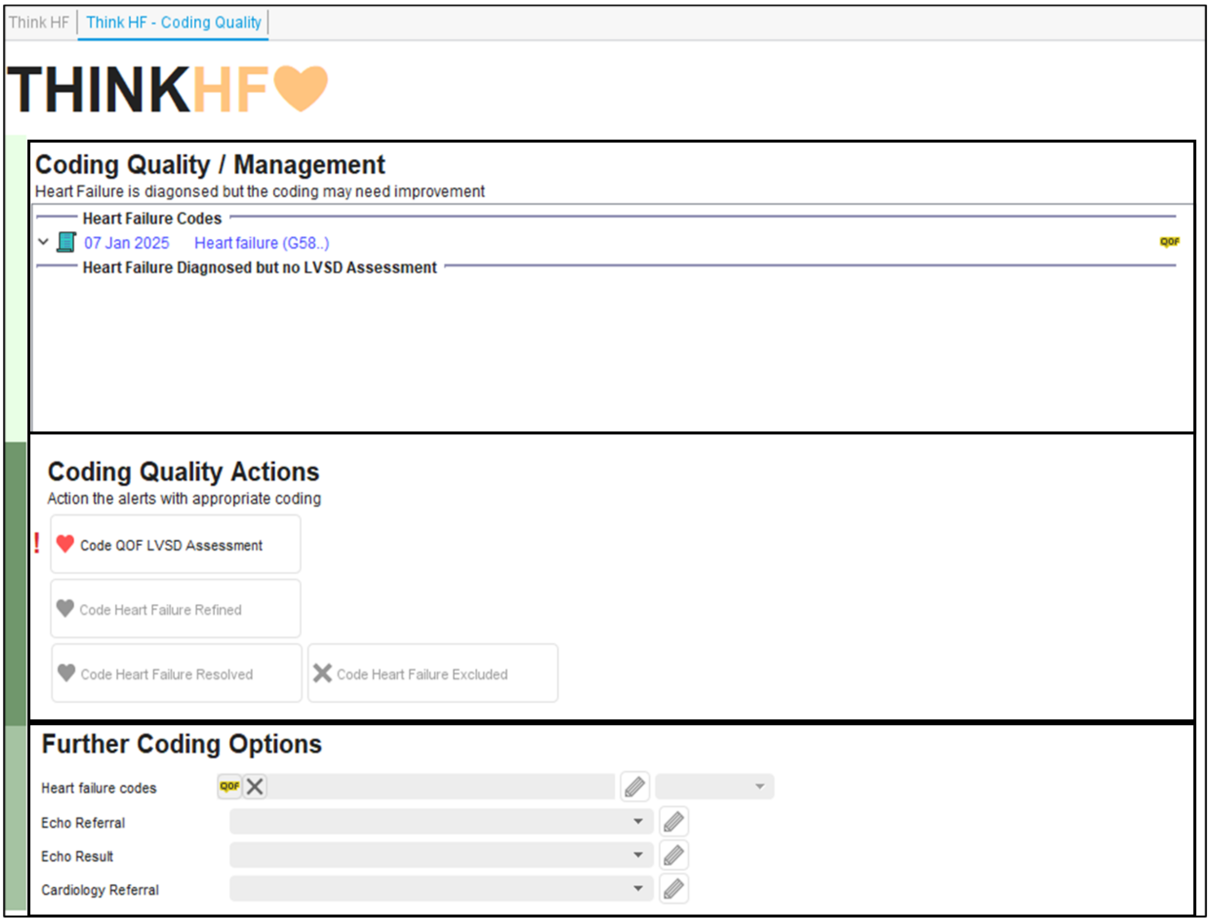
